# Persistent obesity since childhood or adolescence accelerates biological aging in young adults from Chile’s oldest birth cohort

**DOI:** 10.1101/2024.08.14.24311960

**Authors:** mP Correa, R Burrows, C Albala, C Sepúlveda, F Salech, R Troncoso, C Gonzalez-Billault

**Author notes:** **Corresponding should be addressed to:** Dr. María Paulina Correa, Institute of Nutrition and Food Technology, Universidad de Chile. Prof. Christian Gonzalez-Billault, Faculty of Science, Universidad de Chile.

## Abstract

Aging and obesity are primary risk factors for chronic conditions such as hypertension and diabetes. However, a mechanistic correlation between these risk factors has not been fully established. Using a historical birth cohort from Chile, we delve into the relationship between obesity and accelerated aging, spanning cellular to systemic levels. The cohort, comprising men and women in their late 20s, had their BMI recorded since birth, with 57% having obesity since childhood or adolescence. Our aim was to investigate if persistent obesity since childhood or adolescence leads to the display of molecular aging features in young adulthood. We also sought to determine whether cardiometabolic health issues accompanied this early aging phenotype. We used inferential statistics and data mining for analysis. Results show that persistent obesity since childhood or adolescence leads to epigenetic changes, telomere shortening, chronic inflammation, impaired nutrient sensing, mitochondrial stress, and diminished intercellular communication, resembling a compound network of interactions. This obesity-induced accelerated aging phenotype coincided with persistent decline of the cardiometabolic profile. Implications of our findings are significant and suggest that integrating molecular markers with clinical and epidemiological data could be valuable in identifying individuals at increased disease risk due to accelerated aging.

## INTRODUCTION

The likelihood of developing age-related health issues also increases with increasing life expectancy. While normal-weight individuals may develop non-communicable chronic diseases (NCDs) at older ages, overweight or obese individuals often experience the onset of these conditions at a younger age. Although age remains the most significant risk factor for almost all NCDs, obesity also plays a significant role in this epidemic. Extensive research shows that obesity reduces health span by increasing the risk of musculoskeletal, cardiometabolic, and neoplastic abnormalities^[1–5]^. Sarcopenia, atherosclerosis, insulin resistance, and a decline in adaptive immune function are typical in human aging. However, obesity can hasten the initiation and progression of these aging processes into more severe clinical manifestations. Furthermore, these health issues are increasingly seen in younger people^[6–10]^ and may indicate early signs of aging. Recently, two research groups reviewed the then nine hallmarks of aging and the potential interactions between each hallmark and obesity. They both concluded that the pathophysiological changes associated with obesity are similar to or contribute to those seen in aging, suggesting that obesity may hasten the progressive decline in physiological integrity typically observed in aging organisms^[11,12]^. While three aging hallmarks were added in 2023 (chronic inflammation, dysbiosis, and altered macroautophagy)^[13]^, they are also common in obesity, strengthening the hypothesis but still leaving it unproven.

Epidemiological research has consistently linked obesity to shorter lifespan and increased risk of early-onset chronic health disturbances^[14–17]^, but further research is needed to understand the specific molecular pathways and mechanisms connecting obesity and aging. Both share many physiological traits, including dysfunctional adipose tissue, oxidative stress, gut microbiome imbalance, systemic inflammation, shortening telomere length, mitochondrial dysfunction, impaired nutrient sensing, altered intercellular communication, loss of proteostasis^[20–24]^, cellular senescence^[25]^, and age-related DNA hypomethylation in specific cells and tissues^[26,27]^. Understanding this connection at the cellular and molecular levels is crucial for identifying resiliency markers and dynamics that maintain an organism’s health as it ages. Furthermore, studying these common biological markers in a population-based setting is crucial despite the complexities of using humans for aging research (e.g., ethical issues, environmental factors, and limited data on previous exposures). After failing to meet the 2025 WHO goal of stopping the increase in obesity and with an estimated 1 billion people expected to be affected by 2030, we are headed towards a future where the global population may be older than existing sociodemographic data indicates, potentially jeopardizing efforts for healthy, functional, successful aging^[28]^. We must address obesity as a model of accelerated aging in humans if we expect to connect Geroscience with healthcare providers, decision-shapers, and policymakers and pave the way for translational aging research and longevity medicine.

A cohort of males and females born in Chile in the 1990s and exposed to obesogenic environments for a long time could be an ideal setting to test the hypothesis that obesity might accelerate the expression of a phenotype that should appear later in life. At ages 28-29, the mean BMI in the cohort was 29 kg/cm^2^, and 39% had obesity, with no sex differences. Their lipid profile, blood pressure, and pulse wave velocity suggest high cardiovascular risk. Metabolic Syndrome (MetS) and liver disease (NAFLD) increased from 15-24% at 23y to 38-55% at 29y. About 12%-15% of the participants already used glucose-, blood pressure-, or cholesterol-lowering medication as early as 28-29y. These clinical findings suggest that early exposure to obesity has caused these young men and women to age at a faster pace than what is considered physiologically normal. In this study, we will determine whether the presentation of biochemical or physiological aging signatures accompanies this dysfunctional cardiometabolic phenotype and establish whether exposure to obesity since childhood or adolescence contributes to the expression of these aging signatures in young adulthood. Considering the sexual differences in the onset, progression, and incidence of most NCDs, we will also examine the sex-specific features of this obesity-induced accelerated aging phenotype.

## RESULTS

### Study design of the ObAGE project

Between April 2022 and June 2023, we collected blood samples from 205 participants in the Santiago Longitudinal Study (SLS), Chile’s oldest epidemiological birth cohort. The SLS began in 1992 to study the effects of nutrition on children’s growth and health^[9,29,30]^. For the obesity-induced accelerated aging (ObAGE) study, we recruited males and females aged 28-31y who had maintained a healthy BMI throughout their lives (TG1), those who had been persistently obese since adolescence (TG2), and those who had been persistently obese since early childhood (TG3). We used cubic polynomials to analyze the BMI trajectories of SLS participants since birth. Fig. 1 presents the BMI trajectories (Fig. 1A) and health data from previous assessment waves (Fig. 1B-K). We isolated peripheral mononuclear blood cells and obtained genomic DNA using standardized procedures in our labs at Universidad de Chile. We determined epigenetic age using 1st and 2nd-generation clocks at the Clock Foundation (Torrance, CA). We also obtained plasma samples to evaluate glucose and lipid profiles and determine age-related protein content, including cytokines, adipokines, myokines, and growth factors. We conducted comprehensive clinical assessments, including anthropometric and DXA-derived body composition evaluations, blood arterial pressure and pulse-wave velocity measurements, and neck and abdominal ultrasound to evaluate the carotid arteries and the liver, respectively.

**Figure 1.**
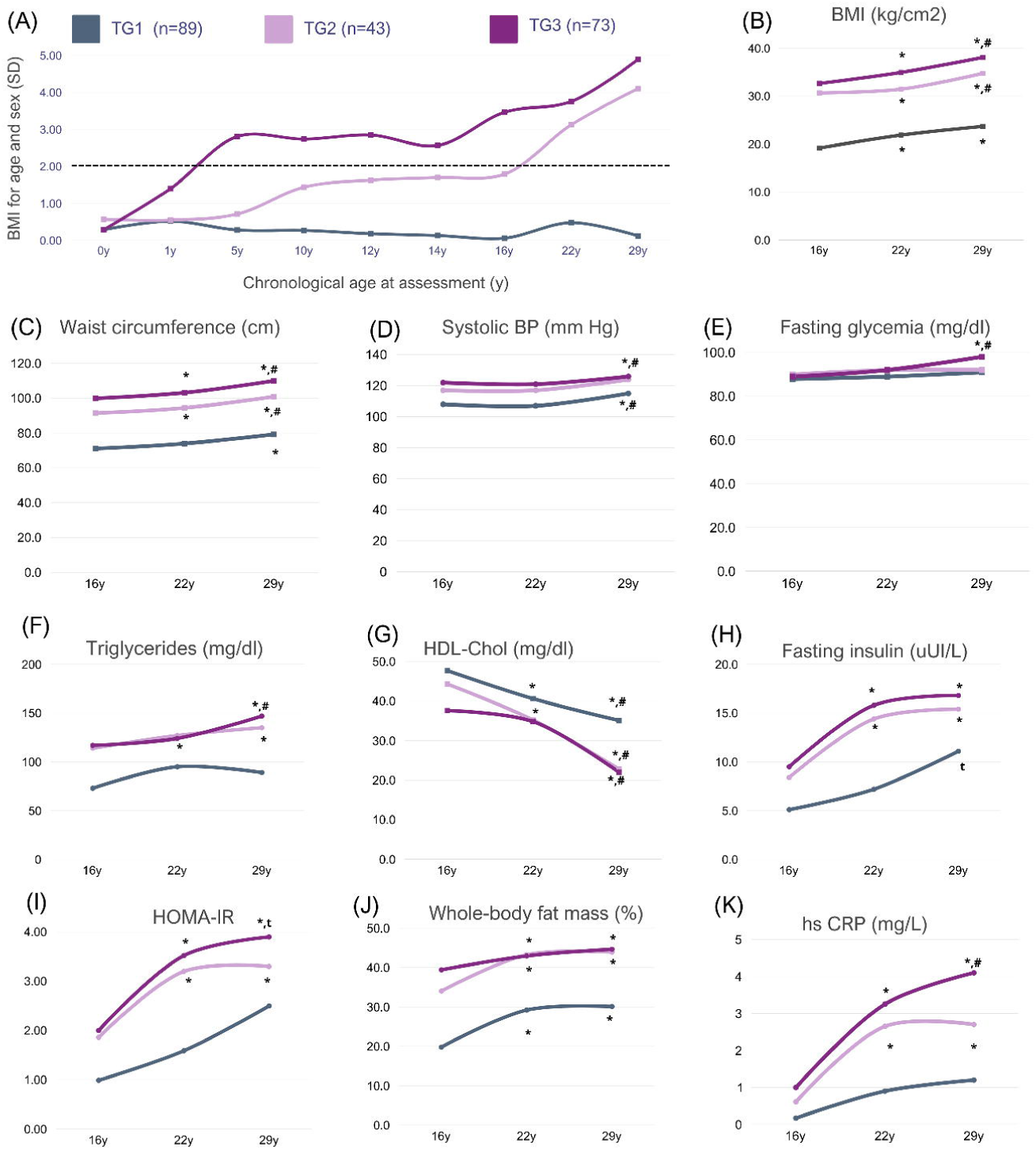
BMI trajectory from birth to adulthood in the study participants and changes in body composition and cardiometabolic markers from adolescence to adulthood. A) Polynomial-based interpolation of BMI trajectory from birth to adulthood. BMI estimated from weight (kg) and height (cm) measured at several time points was standardized (z score) according to the 2007 WHO growth references. Reference values for males and females 19 years and older were used to standardize BMI assessed in adulthood. TG1 refers to participants always having a BMI in the healthy range; TG2 refers to participants with obesity starting in adolescence and remaining obese into adulthood; and TG3 refers to participants who were obese in early childhood and remained obese into adulthood (TG3). (B) to (L) Repeated-measures ANOVA of raw BMI and selected cardiometabolic health biomarkers assessed at 16, 22, and 29 years. (*) Denotes statistical difference from the given biomarker as measured at 16 years. (#) Denotes statistical difference from the given biomarker as measured at 22 years. (t) Denotes trend toward significance from the given biomarker as measured at 22 years. TG1 refers to participants always having a BMI in the healthy range; TG2 refers to participants with obesity starting in adolescence and remaining obese into adulthood; and TG3 refers to participants who were obese in early childhood and remained obese into adulthood (TG3).

### Brief sample description

The study involved 48% female participants. In the sample, 21% had been obese since adolescence and 36% since childhood. There was no link between sex and BMI category. Participants’ ages ranged from 28.0 to 31.3 years, with no differences based on sex or BMI category. Participants became obese around age 2 if it started in early childhood and at 15.8y if it started in adolescence. The average duration of obesity was 12.9y in TG2 and 26.6y in TG3. One male participant had a T2D diagnosis, and three females were taking metformin due to glucose intolerance (doses 500-850 mg/day). A description is available in Supplementary Material (Table S1).

### Significant deterioration of body composition and cardiometabolic markers from adolescence to adulthood in participants with obesity since childhood or adolescence

Repeated measures ANOVA was conducted to examine how BMI, WC, SBP, Gli, Ins, HOMA-IR, TG, HDL, whole-body fat mass (herein FM), and hs-CRP changed over time (16y, 22y, 29y) within each BMI trajectory group (Fig.1B-K; see also Tables S2 and S3 in Supplementary Material). Findings revealed that time notably influenced the changes in body composition and cardiometabolic indicators across all groups during the shift from adolescence to adulthood. A Tukey post hoc adjustment revealed differences worth discussing. At 29, TG1 showed significant increases in BMI, WC, SBP, and FM and decreased HDL compared to their measurements at 16y. Differences in BMI and FM were also observed at age 22 compared to age 16. Furthermore, there were changes in systolic blood pressure and HDL levels between ages 22 and 29. Yet, except for HDL and FM, all markers remained within normal clinical ranges in TG1. In TG2, BMI, WC, TG, HDL, Ins, HOMA-IR, FM, and hs-CRP changed significantly at 29y and 22y compared to 16y. BMI and WC also significantly increased between ages 22 and 29. All markers had worsened at 22y and 30y compared to 16y in TG3, with WC, HDL, FM, and hs-CRP changing significantly between ages 16-22. Notably, at 29y, WC, FM, SBP, Ins, and HOMA-IR levels were above clinical normality, while HDL was well below the recommended levels. Therefore, although time led to changes in the cardiometabolic parameters of the three groups, it imposed a greater health toll on subjects with obesity since childhood and adolescence (OCA).

### Childhood and adolescent obesity significantly impact body composition and cardiometabolic phenotype in adulthood

Anthropometric and cardiometabolic phenotyping differences based on the BMI trajectory group were explored using one-way ANOVA. For anthropometric measurements, the analysis was conducted separately in males and females. We found significant differences in BMI across all categories in males and females, WC in females, and WtHtR in males (Fig.2A-D). In both sexes, FM differed only in TG1 *vs* TG2 and TG3. The same pattern was observed for WtHtR in females and WC in males. Notably, males and females in TG3 exhibited a mean BMI indicating severe obesity, while those in TG2 were just below the 35 kg/cm^2^ cutoff. Both males and females in TG2 and TG3 had WC and WtHtR measurements that were significantly above the limits of clinical normality. Cohen’s *f* coefficient indicated that the effect of life-course BMI trajectory on current anthropometric markers was strong (*f*>0.8) in both sexes.

**Figure 2.**
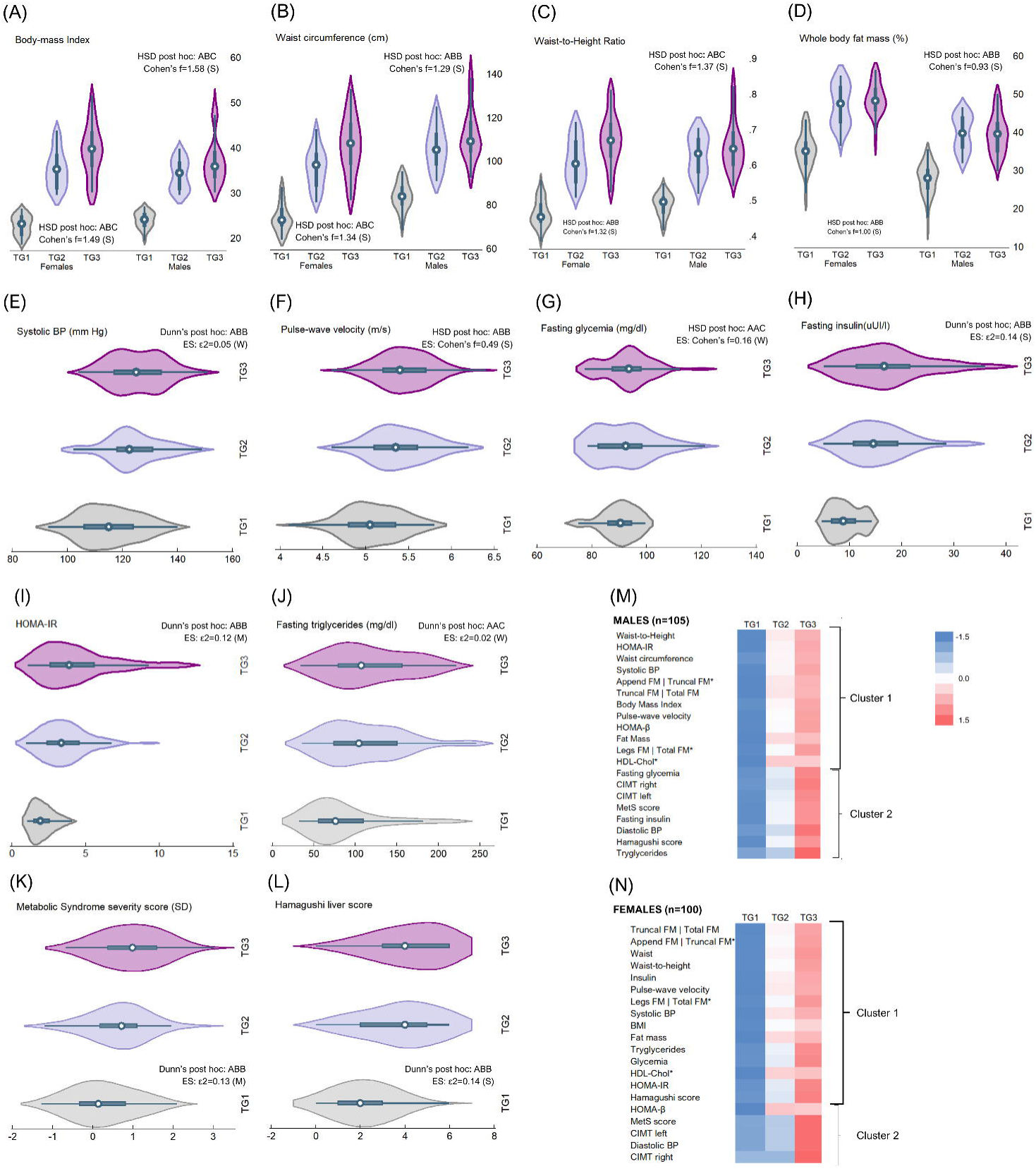
Childhood and adolescent obesity have a significant impact on body composition and cardiometabolic health in adulthood. (A) to (D) Probability distribution, between-group comparison, and effect size for difference (ES) of body composition markers assessed at 28-29y, by life-course BMI trajectory and sex. Data distribution and probability density compared with violin plots. ANOVA with HSD adjustment for between-group comparisons: A=TG1; B=TG2; C=TG3. Different letters indicate significant statistical differences. The same letter denotes means that do not differ. Using Cohen’s f coefficient, ES was determined: 0.10 = weak ES, 0.25 = moderate ES, and 0.40 = strong ES. TG1 refers to participants always having a BMI in the healthy range; TG2 refers to participants with obesity starting in adolescence and remaining obese into adulthood; and TG3 refers to participants who were obese in early childhood and remained obese into adulthood (TG3). (E) to (L) Probability distribution, between-group comparison, and ES of cardiometabolic markers assessed at 28-29y, by life-course BMI. Data distribution and probability density compared with violin plots. ANOVA or Kruskal-Wallis test with HSD or Dunn adjustment was conducted for between-group comparison. A=TG1; B=TG2; C=TG3. Different letters indicate significant statistical differences. The same letter denotes means/medians that do not differ. The effect size for difference (ES) was determined using Cohen’s f (0.10 = weak ES, 0.25 = moderate ES, and 0.40 = strong ES) or epsilon-squared (0.01-< 0.06=weak ES), 0.06 -< 0.14=moderate ES, and >= 0.14=strong ES), depending on data distribution. TG1 refers to participants always having a BMI in the healthy range; TG2 refers to participants with obesity starting in adolescence and remaining obese into adulthood; and TG3 refers to participants who were obese in early childhood and remained obese into adulthood (TG3). (M) Hierarchical cluster analysis was conducted on standardized data of selected anthropometric and cardiometabolic markers assessed at 28-29y, by sex. Heat maps are presented for males and females. Distance criteria: Euclidian method. Linkage criteria: Ward linkage. For this analysis, HDL-chol, legs FM | total FM and appendicular FM | truncal FM are the multiplicative inverse (1/x). TG1 refers to participants always having a BMI in the healthy range; TG2 refers to participants with obesity starting in adolescence and remaining obese into adulthood; and TG3 refers to participants who were obese in early childhood and remained obese into adulthood (TG3).

We used the Kruskal-Wallis test and one-way ANOVA with Dunn and Tukey corrections to examine the differences in the cardiometabolic profile. Our findings showed that TG1 had lower SBP, PWV, Ins, HOMA-IR, MetS severity score, and Hamagushi NAFLD score (Fig. 2E-L) than TG2 and TG3 groups. There were no significant differences between TG2 and TG3 groups. On the other hand, we observed that Gli and TG were higher in TG3 than in TG1 and TG2 groups, with no significant differences between the latter two groups. It is noteworthy that the effect of life-course BMI trajectory on current cardiometabolic markers was weak for SBP, Gli, and TG, moderate for MetS score and HOMA-IR, and strong for PWV, Ins, and Hamagushi NAFLD score, as indicated by Cohen’s *f* coefficient or ε^2^. Although all groups had clinically normal Gli values, the median SBP, Ins, HOMA-IR, and Hamagushi were higher than recommended in TG2 and TG3. MetS severity score was also above risk limits (≥1 SD) in TG3.

We further conducted hierarchical clustering analysis (HCA) to investigate the anthropometric and cardiometabolic profile of the samples and identify sex-specific dysfunctional patterns within this profile. Results are presented as heatmaps (Fig. 2M-N), while phylograms and adjacency matrixes are presented in supplementary material (Fig. S1-S2, Table S4-S5). Except for HOMA-β and CIMTr in females, the heatmaps demonstrate a trend toward values indicating dysfunction or clinical abnormality when transitioning from TG1 to TG2 and TG3 in both males and females. Generally, the highly saturated blue markers denoting values within the clinically normal range (except for HDL) are predominantly found in subjects of TG1, while the red markers, indicating dysfunction, have become more saturated in adult subjects with obesity since childhood. In adult men and women with obesity since adolescence, there is a broader range of shades, from light blue to soft red, suggesting a tendency toward normality in some markers and a tendency toward dysfunction in others. Likewise, phylograms (Fig. S1-S2) revealed two clusters (C) in males and females (Fig. 2M-N, brackets). Of note, C1 included all the anthropometric markers assessed during the clinical appointments in both sexes. Males’ C1 also consisted of SBP, PWV, HOMA-IR, HOMA-β, and HDL, while in females, C1 included all those markers as well as Gli, Ins, TG, and the Hamaguchi liver score. Two major groupings might suggest a similarity between males and females in expressing their anthropometric and cardiometabolic profiles as adults. However, upon closer examination, the within-cluster (C1: 0.025 vs. 0.082 | C2: 0.182 vs. 0.227) and between-cluster (0.412 vs 0.803) variances derived by the HCA reveal that males exhibit lower variations than women. This indicates that obesity since childhood or adolescence has a more consistent effect on the expression of the anthropometric and cardiometabolic profile in adulthood in males compared to females. This confirms that premenopausal women had better protection against obesity-related cardiometabolic changes in adulthood compared to similarly affected men.

### Early expression of aging signatures relates to childhood and adolescent obesity

#### Epigenetic changes and telomeres length

Epigenetic age was estimated using the Human Multi-tissue DNAmAge estimator and DNAm GrimAge. DNAm TL, a biomarker of aging more strongly associated with age than measured TL^[31]^, was also obtained from the same DNA sample. Figure 3A-D shows that the epigenetic age (DNAmAge) of participants in TG2 and TG3 was consistently above their chronological age at the time of the test (CAge). The Pettit test for change-point detection in a trend (Fig. 3A) confirms a shift in the central tendency of the DNAmAge series when transitioning from TG1 to TG2 participants, supporting the hypothesis that OCA may hasten the pace of aging. Of note, in the sample, DNAmAge ranged from 23-43.7y, and DNAmGrimAge ranged from 20.8-41.1y.

**Figure 3.**
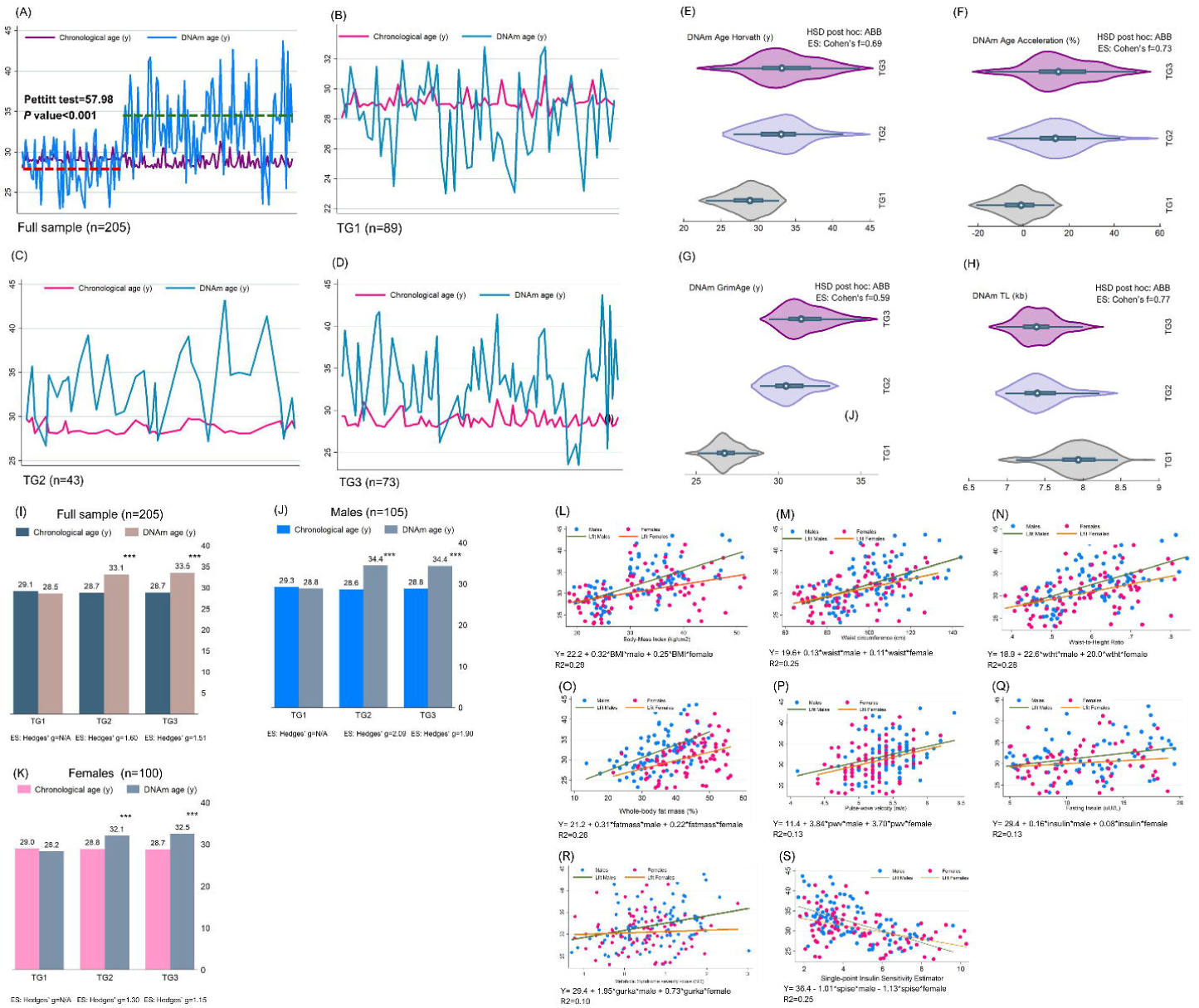
Obesity-related epigenetic changes and telomeres length. (A) to (D) Data visualization of chronological vs. DNAm age in the sample, overall and by BMI trajectory group. In panel (A), Pettitt’s test for homogeneity was employed to detect change points in the data series. TG1 refers to participants always having a BMI in the healthy range; TG2 refers to participants with obesity starting in adolescence and remaining obese into adulthood; and TG3 refers to participants who were obese in early childhood and remained obese into adulthood (TG3). (E) to (H) Probability distribution, between-group comparison, and effect size for difference of body composition markers assessed at 28-29y, by life-course BMI trajectory and sex. Data distribution and probability density compared with violin plots. ANOVA with HSD adjustment for between-group comparisons: A=TG1; B=TG2; C=TG3. Different letters indicate significant statistical differences. The same letter denotes means that do not differ. The effect size for difference (ES) was determined using Cohen’s f (0.10 = weak ES, 0.25 = moderate ES, and 0.40 = strong ES). TG1 refers to participants always having a BMI in the healthy range; TG2 refers to participants with obesity starting in adolescence and remaining obese into adulthood; and TG3 refers to participants who were obese in early childhood and remained obese into adulthood (TG3). (I) to (K) Within-group comparison of chronological age and DNAm age, overall and by sex. Student’s t-test for paired data: * Difference significant at α<0.05; *** Difference significant at α<0.001; t Trend toward significance. The effect size for difference (ES) was determined using Hedges’ *g* for paired comparisons (Gibbons RD, Hedeker DR, Davis JM, 1993. Estimation of effect size from a series of experiments involving paired comparisons. J. Educ. Stat 18, 271–279.): 0.2 = Weak ES; 0.5 = Moderate ES; 0.8 = Strong ES; 1.2 = Very strong ES. TG1 refers to participants always having a BMI in the healthy range; TG2 refers to participants with obesity starting in adolescence and remaining obese into adulthood; and TG3 refers to participants who were obese in early childhood and remained obese into adulthood (TG3). (L) to (S) Multiple linear regression models were used to test whether current sex-adjusted body composition and cardiometabolic markers significantly predict DNAm age.

ANOVA with Tukey corrections examined the between-group differences for these epigenetic-based aging biomarkers (A=TG1; B=TG2; C=TG3; when different letters are used, it indicates significant statistical differences. Therefore, the same letter denotes means that do not differ). As expected, TG1 had lower DNAmAge (28.0y *vs*. 33.2y *vs* 33.5y; ABB), ΔDNAmAge (-3.9% *vs* 15.6% *vs* 16.2%; ABB), DNAmGrimAge (26.5y *vs* 30.9y *vs* 32.0y; ABB), and greater DNAmTL (8.01 kb *vs* 7.46 kb *vs* 7.42 kb; ABB) than TG2 and TG3 groups, with no differences found between TG2 and TG3 (Fig.3E-H; see also Table S6). The pattern holds after accounting for the effect of sex. The effect of BMI trajectory on these epigenetic aging markers ranged from *f*=0.59 (95%CI 0.41-0.77) to *f*=0.77 (95%CI 0.64-0.86), denoting a strong impact of OCA on the epigenetic-based aging profile in adulthood.

A paired t-test compared CAge and DNAmAge within the groups. This was followed by calculating Hedges’s *g* coefficient for paired data as an effect size measure (Fig.3I-K). No differences were found in DNAmAge and CAge for TG1 in the full sample. On the other hand, in both TG2 and TG3, DNAmAge was significantly higher than CAge (MD_TG2_: 4.4y; *p*<.001 | MD_TG3_: 4.8y; *p*<.001). Based on Hedges’ *g* ranging 150-1.60, which denotes a difference of very large size, it can be inferred that 94% of participants in TG2 and TG3 have a DNAmAge higher than the group’s mean CAge. Moreover, there is an 87% chance that a random person picked from TG2 and TG3 will have a DNAmAge higher than their CAge.

To delve deeper into the link between anthropometric and cardiometabolic health markers and DNAmAge, multilevel models were used to also account for sex specificities (Fig.3L-S). The results showed that BMI, WC, WtHtR, and FM could predict from 25% to 29% of DNAmAge variation in adulthood. BMI had the highest predictive ability among all the anthropometric markers, which could have important epidemiological implications. Likewise, PWV, Ins and MetS score predicted from 10% to 13% of DNAmAge variation. Notably, the single-point insulin sensitivity estimator (SPISE), a surrogate measure of IR computed using BMI, TG, and HDL values^[32]^, predicted 25% of DNAmAge variation. Sex-specific regression coefficients were significant for all markers except for PVW and MetS severity scores, where coefficients for females lacked significance.

#### Obesity-related cytokine, adipokines, myokines and growth factors profile

Obesity is characterized by low-grade inflammation, which leads to the release of pro-inflammatory cytokines, human growth factors (GF), abnormal myokines, and other adipokines from adipose tissue. Chronic low-grade inflammation is also a common feature of aging, increasing the risk of premature morbidity and mortality^[40]^. We determined a selection of such biomarkers in ObAGE participants and compared their expression levels between different BMI trajectory groups (Fig.4A-K see also Table S7). For analysis, all variables were log-transformed, except for hs CRP. TG1 had lower expression of hs-CRP (1.96 mg/L *vs*. 3.67mg/L *vs* 4.24mg/L; ABB), IL-2 (0.42 *vs* 0.66 *vs* 0.65; ABB), IL-6 (0.69 *vs* 1.03 *vs* 0.99; ABB), IL-10 (2.18 *vs* 2.27 *vs* 2.36; ABB), FGF-21 (2.21 *vs* 2.42 *vs* 2.45; ABB), and leptin (males 3.81 *vs* 4.28 *vs* 4.31; ABB | females 4.23 *vs* 4.66 *vs* 4.75; ABB) than TG2 and TG3, with no differences found between TG2 and TG3. GDF-15 was higher only in TG3 than TG1 and TG2 (3.44 *vs* 3.45 *vs* 3.51; AAC). No differences between BMI trajectory groups were observed for GDF-11 and TNFα. Of note, Cohen’s *f* values indicate that BMI trajectory strongly affects IL-6 (*f*=0.46), hs CRP (*f*=0.41) and FGF-21 (*f*=0.54) and moderately affects GDF-15 (f=0.31), IL-2 (*f*=0.29), and IL-10 (*f*=0.27) expression in adulthood. BMI trajectory also strongly affected adulthood leptin values in males (f=0.77) and females (*f*=0.75). Additionally, the BMI trajectory strongly affected the expression of IGF-1 (f=0.55) and IGF-2 (f=0.45) during adulthood. The levels of IGF-1 were found to be lower in participants with obesity than TG1, and there were significant differences in all BMI trajectory groups (4.65 vs. 4.55 vs. 4.45; ABC). Additionally, IGF-2 levels were higher in TG1 than in TG2 and TG3, but no differences were found between the latter (5.54 vs. 5.46 vs. 5.44; ABB). It’s important to note that, besides IGF-1, none of the other aging markers assessed in the study showed differences among participants with obesity. Our analysis of log-transformed variables (Fig.4L-S) also shows that the impact of BMI trajectory on myokines expression in adulthood ranged from moderate to strong (f=0.25 to f=0.41). Participants in TG1 had lower apelin (5.49 *vs*. 5.65 *vs* 5.63; ABB), oncostatin (1.59 *vs* 1.72 *vs* 1.70 ABB), log-Myostatin (7.06 *vs* 7.19 *vs* 7.16; ABB), irisin (7.71 *vs* 7.83 *vs* 7.80; ABB), and osteonectin (5.19 *vs* 5.27 *vs* 5.34; ABB), than participants in TG2 and TG3, with no differences found between TG2 and TG3 (Fig.4J-L). Conversely, TG1 had higher Musclin (6.21 *vs* 6.08 *vs* 6.04; ABB). It is worth recalling myokines can be released by both skeletal muscle and adipose tissue^[33]^. They have the potential to act on both, indicating a bidirectional communication between fat and muscle cells. Additionally, myokines play a crucial role in the communication between muscle and other tissues, such as bone tissue, liver, and pancreatic cells. Therefore, they can be key in assessing intercellular communication; altered intercellular signaling has also been regarded as a hallmark of aging, with myokines being identified as markers of aging, muscle weakness, and frailty^[34]^.

**Figure 4.**
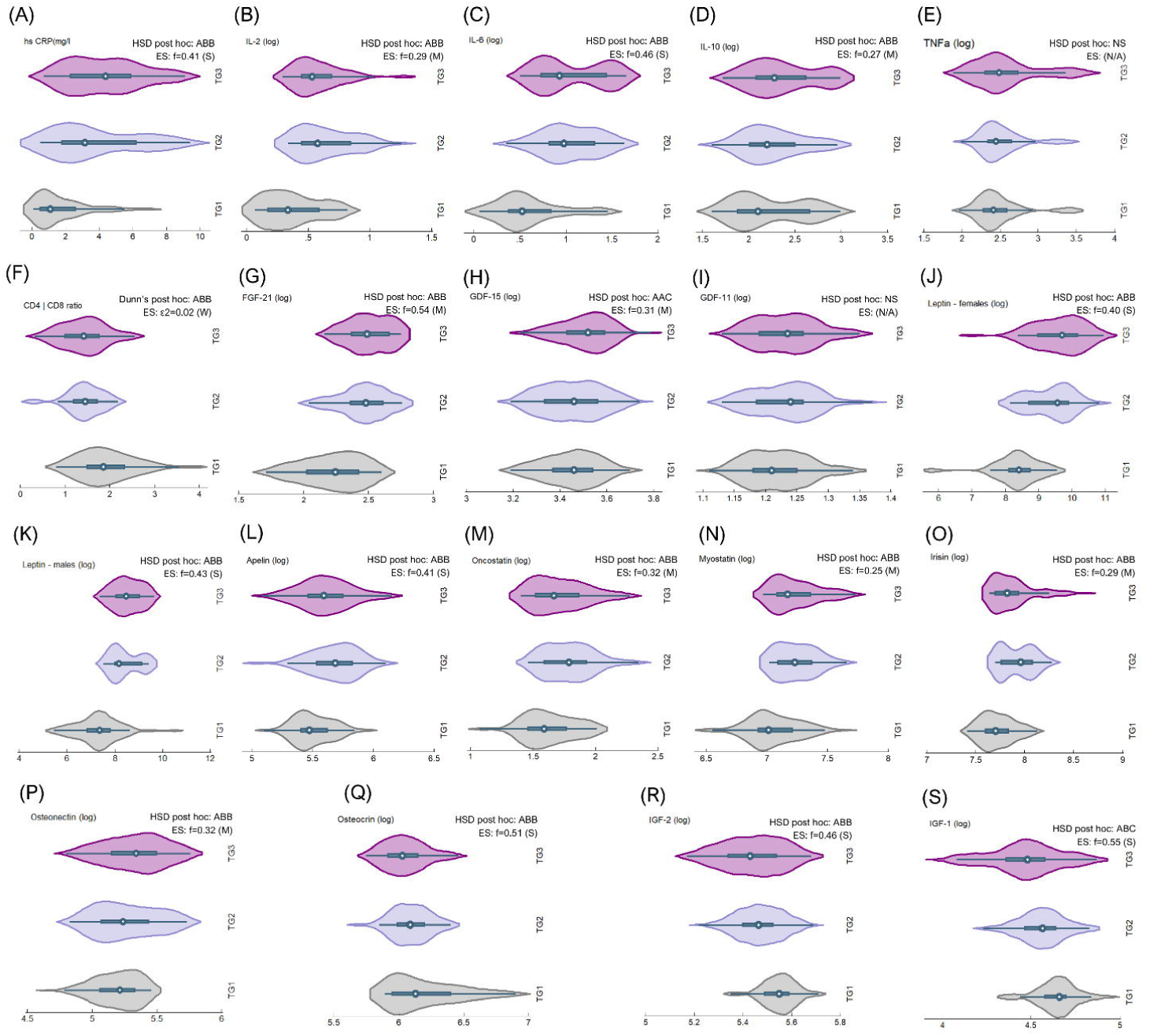
Obesity-related cytokine, adipokines, myokines and growth factors profile. (A) to (S) Probability distribution, between-group comparison, and effect size for the difference of body composition markers assessed at 28-29y, by life-course BMI trajectory and sex. Data distribution and probability density compared with violin plots. ANOVA with HSD adjustment. The effect size for difference (ES) was determined using Cohen’s f (0.10 = weak ES, 0.25 = moderate ES, and 0.40 = strong ES). The CD4|CD8 ratio was compared with KW tests with Dunn adjustment. The ES was measured with epsilon-squared (0.01-< 0.06=weak ES), 0.06 -< 0.14=moderate ES, and >= 0.14=strong ES). TG1 refers to participants always having a BMI in the healthy range; TG2 refers to participants with obesity starting in adolescence and remaining obese into adulthood; and TG3 refers to participants who were obese in early childhood and remained obese into adulthood (TG3).

#### Sex-related features in the expression of obesity-induced accelerated aging phenotypes

We used HCA to study the obesity-induced accelerated aging phenotype expression in both males and females. Because our participants were young, we found that unsupervised learning analysis was more suitable for detecting hidden patterns within the data. Results are shown in Fig. 5 as heatmaps (panels A and D), radial dendrograms (panels B and E), and hierarchical edge bundles, which allow better representation of adjacency relations in compound graphs (panels C and F); adjacency matrices can be found in Supplementary Material (Tables S8-S9). The heatmaps show a trend toward greater expression of aging signatures when transitioning from TG1 to TG2 and TG3 in both males and females. Generally, the highly saturated blue denoting lower aging signs expression is predominantly seen in subjects from TG1, while the red shades, indicating higher expression of aging signs, tend to become more saturated in subjects from TG3. Likewise, we identified two clusters in males and females. They are indicated with brackets in the heatmaps and with colors in the dendrograms (males’ C1: blue, males’ C2: green | females’ C1: orange, females’ C2: purple). The strength of relationships and their hierarchical nature can be best understood through hierarchical edge bundles (panels C and F). In these bundles, deeper connections are represented by more saturated colors. Similarly, it’s easier to observe the connections between lower levels of the hierarchy, highlighting a network-like structure with strong connections among aging hallmarks and the transition from primary to integrative features. In males and females, C1 included markers of DNA hypomethylation and telomere loss in both sexes, which are considered primary aging hallmarks, the leading cause of age-related injury, and the trigger of the damaging cascade that accumulates over time^[35]^. C1 in both sexes also consisted of antagonistic hallmarks. For instance, along with other cytokines and chemokines, upregulated IL-6 and GDF-15 have been used to feature cell senescence *in vitro*, particularly the SASP phenotype^[36]^. Upregulated insulin and downregulated IGF1 and IGF2 may denote impaired nutrient sensing in the developing organism, while upregulated FGF21 and GDF15 have been regarded as markers of mitochondrial stress and possibly dysfunction^[37]^. Lastly, C1 in males and females included markers of systemic inflammation, such as hs-CRP, leptin, IL-6, and IL10, and in females, C1 also added a few markers of altered intercellular communication (Myostatin, Oncostatin, Irisin). Males’ C2 mainly consisted of adipomyokines, a marker of intercellular signaling, while females’ C2 contained markers for systemic inflammation and impaired intercellular crosstalk, both of which have been regarded as integrative hallmarks of aging. When analyzing the composition of the males’ and females’ clusters in panels C and F, we found that C1 displayed more saturated colors than C2. This suggests that in young individuals exposed to OCA, the primary and antagonistic hallmarks are expressed more prominently and consistently than the integrative hallmarks, consistent with the idea of a hierarchy between hallmarks of aging.

**Figure 5.**
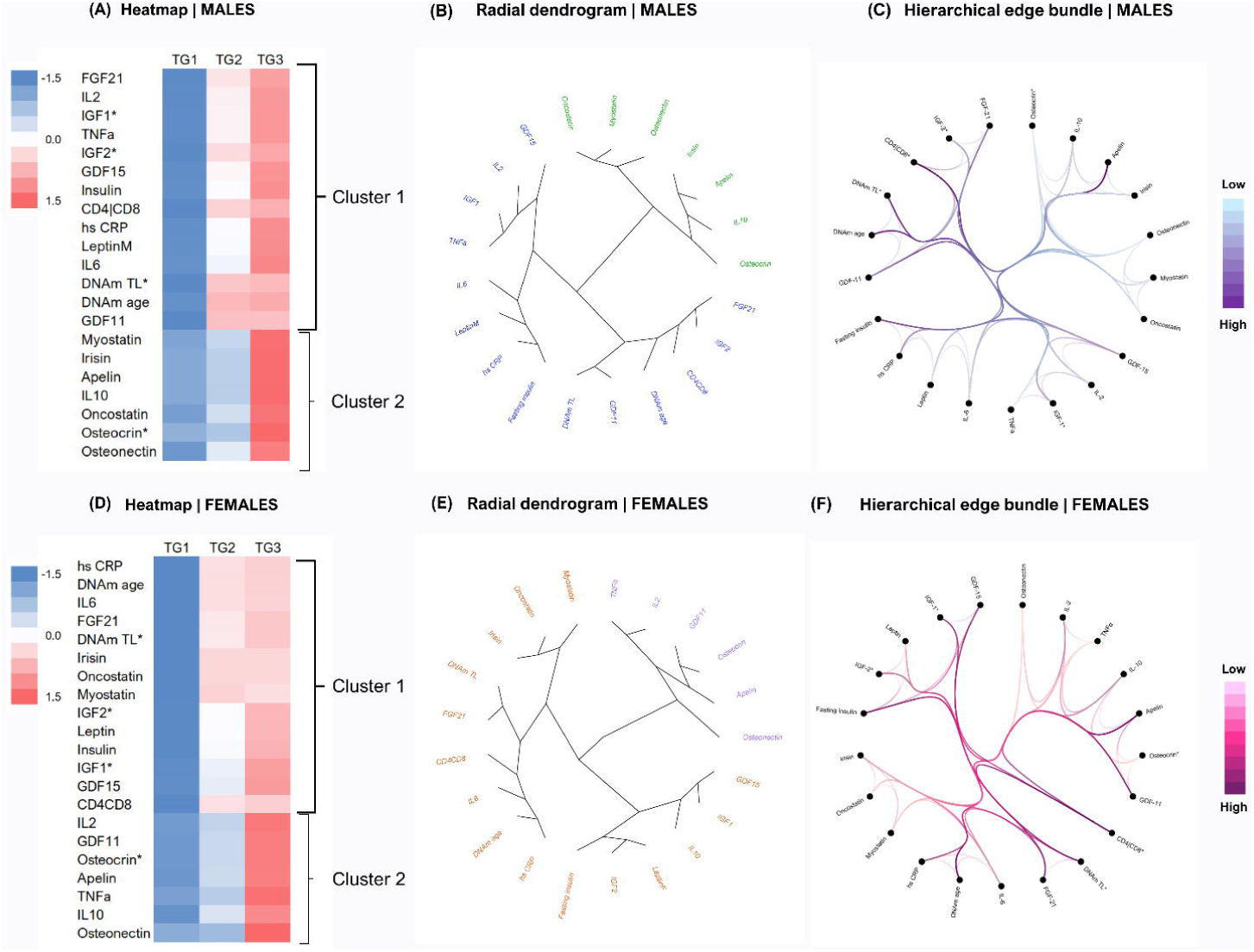
Sex-related features in the expression of obesity-induced accelerated aging phenotypes. (A) to (E) Hierarchical cluster analysis conducted on standardized data of selected aging analytes assessed at 28-29y, by sex. Heatmaps (panels A and D), radial dendrogram (panels B and E), and hierarchical edge bundles (panels C and F) are presented for males and females. Distance criteria: Euclidian method. Linkage criteria: Ward linkage. DNAmTL, IGF1, IGF2, Osteocrin (Musclin), and CD4|CD8 ratio were expressed as the multiplicative inverse (1/x) for analysis. TG1 refers to participants always having a BMI in the healthy range; TG2 refers to participants with obesity starting in adolescence and remaining obese into adulthood; and TG3 refers to participants who were obese in early childhood and remained obese into adulthood (TG3). In panels B and E, different colors represent different clusters (Males’ C1: blue; Males’ C2: green; Females’ C1: orange; Females’ C2: purple). In panels C and F, the more intense color of the trace indicates closeness in hierarchy or similarity in analyte behavior.

### Epigenetic age in adulthood can be predicted from historic repeated BMI measurements

To further explore the contribution of BMI across the life course on biological aging in adulthood, we used repeated measures of standardized BMI (BAZ) to predict DNAmAge and DNAmGrimAge. While there is limited research on analyzing repeatedly measured independent variables, it can be particularly useful for constructing prediction models related to biological aging, given the cumulative nature of the pathophysiology of age-related NCDs. The use of routine clinical markers with repeated measurements can be especially interesting for constructing such models, and this approach can lead to significant advancements in bringing Geroscience to population-based health research. We relied on Welten *et al*. to model a repeatedly measured independent variable (BAZ) and a long-term fixed outcome (epigenetic age) into a prediction model^[38]^. Multilevel modeling tested if BAZ collected from birth to 28-29y significantly predicted epigenetic age, accounting for within-and between-group variations (Fig. 6 A-B). The fitted regression model was: DNAmAge = 1.99 + 0.60*TG1*(mBAZ) + 4.30*TG2(mBAZ) + 2.62*TG3*(mBAZ) + 0.90*(age*male) + 0.86*(age*female) -10.7*(BAZgrowthtrend) + 9.1*(BAZgrowthtrend^2). The overall regression was statistically significant (R-sq_overall_=0.45, R-sq_within_=0.18, R-sq_between_=1.00; Wald (7,21)=135.5, *p*<.0001), and the residuals were randomly scattered around the horizontal axis, indicating the model is a good fit (Fig. 6 C-D). BAZ significantly predicted DNAmAge in TG3 (β=4.30, *p*<.0001) and TG2 (β=2.62, *p*<.0001). However, it did not significantly predict DNAmAge in TG1 (β=0.60, *p*=0.42). When DNAmGrimAge was the predicted variable, we observed the same patterns, though lower overall goodness-of-fit (R-sq_overall_=0.32, R-sq_within_=0.05, R-sq_between_=1.00). According to our results, for a one-unit increase in BAZ during the life course, DNAmAge in adulthood increases by 4.3y and 2.6y in TG2 and TG3, respectively. Next, using the predictive model of epigenetic age, we computed DNAmAge in our sample and compared the results with those determined in DNA samples. We found no significant difference between determined *vs* predicted DNAmAge overall in the sample and after controlling for sex and BMI trajectory group (Fig.6 E-H).

**Figure 6.**
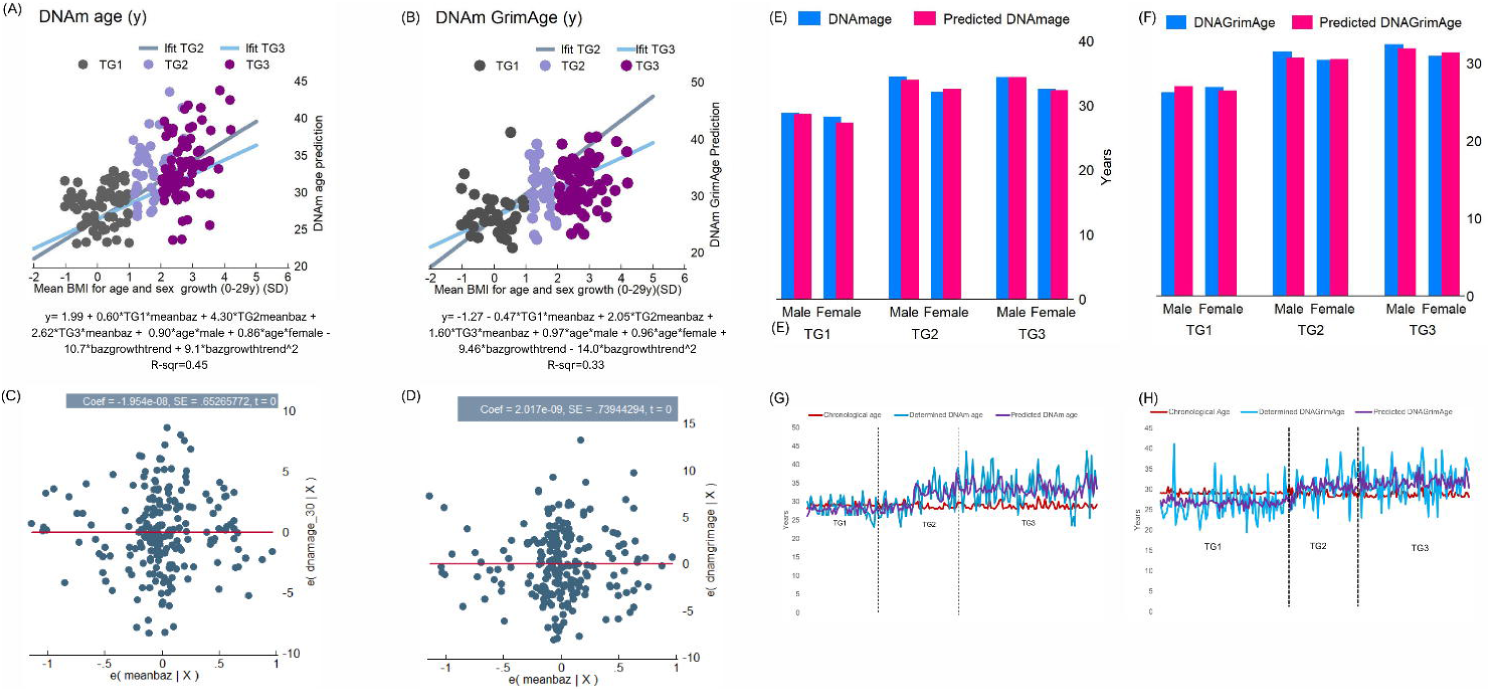
Epigenetic age in adulthood can be predicted from historic repeated BMI measurements. (A) and (B) Prediction equation of epigenetic age using repeated measures of BMI for age (BAZ) and sex from 0-29y. Models were adjusted for BMI trajectory group, sex, age, and BMI growth trend. To create the prediction model, we used Welten et al.’s approach to model a repeatedly measured independent variable (BAZ) and a long-term fixed outcome (epigenetic age). (C) and (D) Residual plots for the prediction models of epigenetic age using repeated measures of BMI for age (BAZ) and sex from 0-29y. (E) and (F) Mean comparison of determined and predicted DNAmAge and DNAGrimAge values after controlling for sex and BMI trajectory group (Student’s t-test for non-independent data). (G) Data visualization of chronological age determined and predicted DNAm age in the sample. (H) Data visualization of chronological age determined and predicted DNAGrimAge in the sample.

## DISCUSSION

The harmful effects of biological aging begin with dysfunction and progress to damage, disease, disability, and ultimately death, impacting cells, tissues, and organ systems. Geriatrics Medicine primarily focuses on older people and deals with the latter four stages. However, dysfunction begins much earlier in life and sets the stage for the next phases. Targeting dysfunction is the most promising way to slow down or postpone the subsequent steps. Obesity, which is highly prevalent in people of all ages, is a significant cause of biological dysfunction, potentially triggering the aging process^[11,12,39]^. In this study, we conducted thorough clinical, physiological, and biochemical evaluations on individuals in their late 20s from Chile’s oldest epidemiological birth cohort aiming to detect early expression of molecular aging biomarkers and establish a link suggesting a causal relationship with obesity persisting from childhood and adolescence into adulthood (OCA). Additionally, we sought to establish a link between this early aging phenotype and obesity-related cardiometabolic dysfunction. This study is unique in that it is the first to test the hypothesis of obesity as a driver of early-onset aging in young adults from a longitudinal birth cohort, assessing a comprehensive panel of markers spanning from cells to systems and accounting for exposures in crucial developmental periods. It also creates synergies between NCD epidemiology, molecular biology, and geroscience. Our findings suggest that OCA triggers premature physiological decline, leading to the expression of molecular aging signatures as early as 28-31y. These signs include a 14-17% increase in epigenetic age compared to chronological age, with some individuals showing up to a 48% increase, as well as telomere attrition, chronic inflammation, and compromised intercellular communication.

The link between increased BMI and accelerated epigenetic aging was initially reported by Horvath et al. They observed that for every 10-point increase in BMI, the age of liver cells was predicted to increase by 2.7y^[26]^. While Horvath did not find a connection between BMI and the epigenetic aging of visceral adipose tissue (VAT), a study of Canadian patients with severe obesity did find higher epigenetic age in the VAT of these individuals compared to VAT from normal-weight controls^[27]^. A meta-analysis of biological, environmental, and social factors influencing epigenetic aging revealed that BMI was linked to accelerated epigenetic aging according to the Horvath, Hannum, Levine, and GrimAge clocks, and for some clocks, was indeed the greatest contributor^[40]^. Predicting up to 20% of BMI variance from age-related DNA methylation was also feasible, which aligns with our findings. Our own research supports the evidence and, for the first time, reveals the presence and extent of these connections in a Hispanic population, an underrepresented group in aging research. In our sample, whole-body fat mass and BMI could predict 26% and 29% of the DNAmAge variance, respectively. Additionally, historical BMI data collected from birth to adulthood allowed for the prediction of 45% of the variability of the epigenetic age at assessment, suggesting a long-term relationship between BMI and epigenetic age.

Another significant finding is the substantial impact of OCA on the signs of aging in adulthood. The impact on most assessed biomarkers, as measured by the effect size for the difference, ranged from strong to moderate. Overall, all epigenetic-age-related biomarkers were found to be strongly affected by persistent obesity since childhood or adolescence. Notably, the impact of OCA on TL in adulthood was the strongest among all biomarkers (f=0.77), despite most studies finding a tendency towards an inverse correlation between TL and obesity^[41]^. It is important to remember that epigenetic changes and telomere length attrition are primary aging hallmarks, the root causes of cell and tissue damage. Although our findings do not conclusively indicate which biochemical signs of aging appear first in individuals exposed to OCA, it is evident that long exposure to this pathological condition has a greater impact on primary aging hallmarks than antagonistic and integrative hallmarks. Therefore, we have initial evidence that obesity may accelerate aging by affecting the molecular responses that initiate damage.

Also noteworthy is the effect size for the difference in two components of the insulin-like growth factor family, IGF1 and IGF2, and the direction of their relationship with the BMI trajectory. Based on Cohen’s *f* values, IGF2 was the non-epigenetic biomarker of aging most affected by OCA. Participants suffering from this condition show decreased values of IGF2 in adulthood. Decreased expression of this growth factor has been linked to the aging of various organs and primordial germ cells. It is postulated that reduced IGF2 compromises the functionality of the mitochondria^[42,43]^. IGF1 was also found to be reduced in participants with OCA. Earlier studies suggested a link between high IGF-1 levels and negative outcomes related to mortality and age-related health issues^[44]^. However, subsequent research has shown that the advantages of lower IGF-1 are more visible in older individuals. In their analysis of data from the UK Biobank from individuals in various age groups, Zhang et al. discovered that IGF1 is a non-linear risk indicator that interacts with age to alter the risk for various health outcomes. Specifically, elevated IGF-1 is mainly linked to reduced disease risk in younger individuals^[45]^. Conversely, it is associated with higher illness and death rates in older individuals^[46]^. Therefore, lower IGF1 levels in participants with OCA in our sample might suggest a higher risk of health issues consistent with an altered cardiometabolic profile. According to some reports, a possible explanation is that IGF-1 levels decrease with age, so higher IGF1 in young adulthood can be considered a youth biomarker^[45,47]^. Diminished IGF1 levels in the elderly lead to loss of resiliency, while IGF1 is crucial for normal development in younger individuals as the focus is on growth and expansion during youth rather than preservation^[47]^.

OCA significantly impacted proteins such as hsCRP, IL6, and leptin and had a moderate-to-strong effect on IL2 and IL10, all well-known biomarkers of systemic inflammation. Conversely, it did not affect the expression of TNFα and GDF-11. GDF-11 typically decreases with age, but it was also found to remain unchanged in individuals with obesity and T2D^[48]^. TNFα plays a crucial role in the link between obesity and diabetes. It may be necessary to have severe insulin resistance for TNFα to be significantly upregulated in individuals with obesity, especially if they are young. Systemic inflammation is a newly recognized hallmark of aging, stemming from genomic instability, epigenetic dysregulation, impaired proteostasis, insufficient autophagy, or the buildup of senescent cells^[9]^. Inflammation, in turn, paves the way for other signs of aging. For instance, it plays a significant role in intercellular signaling and the immune system’s ability to combat age-related damage. In our sample, participants with OCA presented dysregulated values of several myokines (Apelin, Irisin, Myostatin, Oncostatin, Musclin, and Osteonectin), a group of proteins able to mediate muscle–organ crosstalk to the brain, adipose tissue, bone, liver, gut, pancreas, vascular bed, and skin^[49,50]^. Obesity and aging are associated with dysregulated myokine secretion and signaling, contributing to skeletal muscle loss and metabolic disturbances. Thus, it should not be surprising that these markers of intercellular communication are increased in obese subjects compared to normal weight controls, particularly apelin and irisin, which tend to decline in the aging organism. In a developing organism, such as our participants, the increase in these myokines may be a response to metabolic dysfunction induced by obesity. This upregulation could be a reaction to improve insulin sensitivity in obese individuals, or it could be the consequence of reduced sensitivity to its effects, as seen with insulin and leptin in obesity^[51,52]^. Also, systemic inflammation relates to dysfunctional innate and adaptive immunity, a condition known as immunosenescence^[53]^. In our sample, OCA affected a marker denoting a weaker immune system response, such as the CD4+/CD8+ ratio, which was significantly reduced in individuals with persistent obesity. Although the effect size for the difference was small in this case, this response may result from sustained overexpression of proinflammatory cytokines and might indicate the early onset of immune dysfunction. At the systemic levels, inflammation is associated with several health issues, including endothelial dysfunction, insulin resistance, hepatic steatosis, and metabolic syndrome. In our study, individuals with OCA exhibited markers of these alterations, such as elevated values of SBP, DBP, and PWV; insulin, HOMA-IR, and HOMA-β; Hamagushi liver score; and MetS severity score. All these changes are the precursor to age-related diseases; thus, it is reasonable to assume that just as older individuals exhibit frailty as a sign of aging, younger individuals may exhibit dysfunction and cardiometabolic damage as initial signs of aging.

With very few exceptions, the signs of aging in our sample were expressed similarly in participants with obesity, regardless of onset in childhood or adolescence. This supports the notion that persistency, rather than the onset time, is the key factor in obesity-related dysfunctions^[30,54,55]^. Whether this uniformity persists as these young adults approach middle or old age remains to be investigated. However, it appears that the progression to disease is clear and ongoing, as dysfunctional cardiometabolic biomarkers in individuals with OCA fell outside healthy ranges in most cases. Also, IR, NAFLD, and MetS affect 70%, 34%, and 66% of participants with OCA. The preeminence of dysfunction and damage over disease at this stage is unsurprising. It indicates there is still enough resilience to counteract dysfunction and damage from progressing to disease, a trait possibly related to participants’ young age. Moreover, there might be untapped potential to boost this resilience through lifestyle adjustments or pharmacological treatments. If obesity can be used as a model for accelerated aging, it could provide an opportunity for translational aging research and clinical trials of anti-aging interventions. Current ethical regulations restrict the recruitment of healthy older adults for such trials due to higher risks than benefits for healthy research subjects. Additionally, it is difficult to classify a physiological process affecting all living organisms as a disease^[56]^. However, these restrictions do not apply to obesity, especially in younger individuals displaying signs of aging and cardiometabolic damage, for whom disease prevention is crucial.

The presence of molecular aging markers coupled with a dysfunctional cardiometabolic profile in developing organisms strongly supports the concept that health encompasses a set of organizational and dynamic characteristics that maintain physiology rather than simply denoting the absence of pathology^[57]^. For instance, in our sample, only one subject had T2D diagnosis, while 70% showed impaired insulin sensitivity. Only three of those with impaired insulin sensitivity were using metformin. Aside from obesity, our cohort could be considered healthy if health is defined as the lack of disease. However, delving into the cell and molecular levels reveals a markedly different reality, where homeostatic maintenance seems to exist in a weak balance. Future studies should explore how long this balance can be sustained and what preventative or therapeutic actions can delay the transition to age-related multimorbidity. It is crucial to investigate these questions as disease-free young adults, who will reach the prime of middle adulthood within the next 10-15 years, are already displaying clear signs of age-related physiological disruption.

We made noteworthy findings in our investigation of how aging-related molecular markers manifest in both males and females. The aging markers in males and females with obesity generally follow similar patterns. However, some differences worth commenting on are observed in those who have been obese since adolescence. Males in TG2 appear to have better resilience in regulating the expression of myokines, possibly due to their higher muscle mass compared to females. On the other hand, women in this group show less increase in certain chronic inflammation markers, possibly because some of their fatty tissue deposits serve a physiological function, especially considering their fertile age. However, this resilience is no longer found in TG3 women. Likewise, we found that the ObAGE phenotype exhibited strong connections and dynamic interactions among these molecular features in both sexes, resembling a multilevel or compound network. This entanglement aligns with the concept that the hallmarks of biological aging form a complex and dynamic network of interactions, with a bidirectional non-separability between the parts and the whole^[35,39]^. Confirmation of this complex system pattern will require adding more aging biomarkers to our analysis, although some authors argue that the key properties of this network can be measured simply using a small number of biomarkers^[58]^. Further investigation will also be needed to understand how this compound network evolves as these young men and women reach middle and older adulthood and to explore the impact of varying durations of obesity. While the shift to a complex systems perspective on aging has already been suggested, it is still pending ^[59,60]^. Access to large-scale genomics and phenomics data, synergies between epidemiological settings and advanced molecular biology, and new stochastic-based analytical tools should make this transition easier.

We found a few subjects (n=7) who, despite having OCA, had both DNAmAge and DNAmGrimAge well below their CAge; the gap ranged from -3.9% to -16.7% of CAge, according to the Horvath clock. We also found that these individuals do not fit the criteria for metabolically healthy obesity, as they do have cardiometabolic disturbances. Interestingly, the expression of these issues does not follow a specific pattern (see Table S10-S12 and Fig. S3-S4). These resilient individuals include six women, five TG3 participants, and one subject under metformin, which might be a confounding factor. Details of their main features can be found in the supplementary material. Of note, the expression of IGF-1 and IGF-2 was more in line with the mean values observed in TG1 participants (Table S13 and Fig. S5). It is unclear whether the expression of growth factors precedes the stability of the epigenome or if the absence of age-associated epigenetic changes leads to the stability of the insulin-like growth factor system. Determining the exact causal network is a challenging task for future research. Likewise, whole genome sequencing analysis may offer more insights into the ability of these young individuals to effectively manage physiological damage despite experiencing obesity from an early age. Considering the sex distribution, it is worth investigating the potential sexual dimorphism in the expression of this resilience profile.

In conclusion, obesity during important developmental stages, like childhood or adolescence, leads to the display of aging signals such as epigenetic changes, shortened telomeres, systemic inflammation, impaired immune function, disrupted nutrient regulation, and compromised intercellular communication in individuals in their late 20s. Even in young adults exposed to a harmful influence during developmental stages, this so-called ObAGE phenotype manifested through a strong interconnectivity of molecular aging signatures. Furthermore, the ObAGE phenotype is also linked to significant cardiometabolic issues, potentially leading to the early development of cardiometabolic diseases. One significant research challenge will be to determine the timeframe in which cardiometabolic dysfunction progresses into a disease in individuals with the ObAGE phenotype. This is crucial because being diagnosed with a cardiometabolic disease significantly increases the risk of multimorbidity. While this is well established for individuals in middle adulthood, it is yet to be seen to what extent this also occurs in relatively young adults. Considering that the ObAGE phenotype significantly impacts the epigenome and that the study participants are in their prime reproductive years, an important avenue for research will be to investigate whether this phenotype can be passed down to offspring and, if so, whether the effects are consistent when inherited from either the mother or father. Additionally, it will be crucial to determine whether the phenotype manifests differently in male and female offspring.

## Abbreviations

BAZ: BMI for age and sex
BMI: Body mass index
CAge: Chronological age
CIMT*l*: Carotid intima-media thickness (left)
CMIT*r*: Carotid intima-media thickness (right)
DBP: Diastolic blood pressure
DNAmTL: DNA methylation estimated telomere length
DXA: Dual-energy X-ray absorptiometry
FGF21: Fibroblast growth factor 21
FM: Whole body fat mass
GDF11: Growth Differentiation Factor 11
GDF15: Growth Differentiation Factor 15
Gli: Fasting glycemia
HCA: Hierarchical clustering analysis
HDL: High-density lipoprotein
HOMA-IR: Homeostatic Model Assessment -Insulin Resistance
HOMA-β: Homeostatic Model Assessment -β cell function
hs CRP: High sensitivity C reactive protein
IL10: Interleukin 10
IL2: Interleukin 2
IL6: Interleukin 6
Ins: Fasting insulin
MetS: Metabolic Syndrome
NAFLD: Non-alcoholic fatty liver disease
NCDs: Non-communicable chronic diseases
OCA: Obesity since childhood or adolescence
PWV: Pulse wave velocity
SBP: Systolic blood pressure
SPISE: Single-point insulin sensitivity estimator
T2D: Type-2 diabetes
TG: Fasting triglycerides
TNAα: Tumoral Necrosis Factor α
VAT: Visceral adipose tissue
WC: Waist circumference
WtHtR: Waist-to-height ratio

## Data Availability

All data produced in the present study are available upon reasonable request to the authors.

## METHODS

### Ethics declaration

Ethical approval was obtained from the Scientific Ethics Committee at the Institute of Nutrition & Food Technology (INTA), Universidad de Chile. (resolution date: December 1, 2021; resolution number: 21–037). Informed and written consent was obtained from all participants included in the study according to the norms for Human Experimentation^[1]^.

### The SLS cohort

Our research is based on the Santiago Longitudinal Study (SLS), a longitudinal prospective cohort of approximately 1000 Chileans aged 30-31 years, 50% of whom are females. The study participants belong to low-to middle-socioeconomic backgrounds and were involved in research related to nutrition and development as infants. The participants were enrolled at birth and have been followed up at ages 1, 5, 10, 12, 14, 16, 21, and 23 years. They were born in 1992-1996, at term, of uncomplicated vaginal births, weighed >3.0 kg, and were free of acute or chronic health problems^[2]^. Enrollment occurred just as the country entered a period of rapid socioeconomic changes, which saw a decrease in undernutrition and infectious disease and an increase in overnutrition and NCD^[3-4]^. In 2009, when the participants were 16, they were invited to participate in an NIH-funded study to identify the biopsychosocial factors contributing to adolescent obesity and cardiovascular risk. The anthropometric and cardiometabolic markers were assessed again at age 23, and a third wave of assessment is currently ongoing. It is worth mentioning that SLS participants were born during a dramatic nutritional transition from parents or grandparents who were likely exposed to protein-calorie undernutrition, resulting in higher cardiometabolic risk (thrifty phenotype).

### The ObAGE study

The Obesity-induced accelerated aging (ObAGE) Study was conceived as a multiple events case-control (MECC) study embedded in a prospective cohort. In the MECC design, there is a known and enumerated cohort, from which subjects are selected for additional measurements. Sun et al. demonstrated that MECC is superior to case-cohort or nested case-control studies as the former eliminates bias and improves the efficiency of the same data analysis^[5]^. We included participants with complete data in all assessment waves who fell into one of the following categories of lifetime BMI trajectory: (1) participants always having a BMI in the healthy range (TG1); (2) participants with obesity starting in adolescence and remained obese into adulthood (TG2); (3) participants who were obese in early childhood and remained obese into adulthood (TG3). ObAGE plans to enroll n=300 participants, though we relied on a sample of n=205 for this analysis. This sample size allows for testing the study hypotheses and detecting moderate to small differences (f=0.15) at α=0.05 and 1-β=0.8. A full description of the ObAGE study protocol is in Correa et al.^[6]^.

### Anthropometric and body composition profiling

#### At current assessment-wave

The research staff followed standardized procedures to measure weight and height of the participants. Weight was measured to the nearest 0.1 kg, using a precise electronic scale (Seca 703, Seca GmbH & co. Hamburg, Germany) and height was measured to the nearest 0.1 cm, using a Holtain stadiometer with the head in the Frankfurt position. Waist circumference was measured with a non-elastic flexible tape at the midpoint between the edge of the last rib and the iliac crest. The measurements were recorded at 0.1 cm (Seca 201, Seca GmbH & co. Hamburg, Germany). The measurements were taken twice, and a third measurement was taken if the difference between the first two measures exceeded 0.2 kg in weight, and 0.5 cm in height and waist circumference. Body-Mass Index was estimated. A body composition analysis was conducted using a dual-energy X-ray absorptiometry (DXA) machine (Lunar Prodigy Corp., Madison, General Electric, WI), along with Lunar iDXA ENCORE 2011 software (Version 13.60.033 Copyright © 1998-2010). The scan was performed in the fasting state to determine the amount and percentage of body fat and lean mass in various body parts such as arms, legs, trunk, and the total body.

#### BMI estimation in previous assessment waves and modelling of BMI trajectory

BMI estimated from weight (kg) and height (cm) measured at several time points from birth to adulthood was standardized (*z* score) according to the 2007 WHO growth references^[7]^. Reference values for males and females 19 years and older were used to standardize BMI assessed in adulthood. We used a cubic polynomial spline to interpolate each participant’s BMI trajectory from birth to early adulthood. This method takes the data points from original measurements, and the splines smooth the transition between data points. Spline interpolation is often favored over other polynomial interpolation methods (such as Lagrange and Newton polynomials) because it can be used for both segments and entire data series. It also allows for small interpolation errors even when using low-degree polynomials for the spline^[8]^. The second advantage of this method is that it allows the construction of a smooth and visually pleasing parametric curve when dealing with sparse data^[8]^, particularly if the spline departs from the original data points^[9]^, as was the case here (we had data for birthweight in all participants). Thus, spline interpolation is more consistent with how BMI changes over time than linear approaches, in which two data points are connected through a straight line. Linear interpolation uses a linear function for each interval, and although quick and easy, it is not very precise. On the other hand, Spline interpolation uses low-degree polynomials in each interval and chooses the polynomial pieces to fit smoothly together^[8,9]^. Following the same method described in Correa et al. ^[10]^, we fitted models with data measured at birth, 1 year, 5 years, 10 years, 12 years, 14 years, 16 years, 21 years, 23 years, and 28-29 years and obtained individual BMI trajectories from birth to adulthood. Using the full BMI trajectories available, we estimated the timing of obesity onset and duration in those participants who ever had obesity, with a precision of weeks. Python 3.0 was used for data interpolation and BMI trajectory modeling.

### Cardiometabolic phenotyping

#### Blood pressure and pulse wave velocity

Three blood pressure (BP) measurements were made, with the participant at rest seated for at least 15 minutes. An individual was defined as being normotensive when presenting with systolic blood pressure (SBP) < 120 mmHg and diastolic blood pressure (DBP) < 85 mmHg. In all cases, BP was measured at the upper arm using an OMRON 705IT oscillometric monitor following international and national guidelines^[11]^. Aortic pulse-wave (PWV)(m/s^2^) velocity, one of the most important clinical parameters for evaluating cardiovascular risk and vascular adaptation, was measured with an oscillometric technique using the Arteriograph system (TecnoMed, Madrid, Spain). This device measures the pulse wave at a single location using a brachial cuff inflated to 35 mmHg over the SBP. The time between the first systolic wave and the second reflected wave is measured as transit time. To calculate pulse wave velocity, the device uses the ratio of the traveled distance between the jugulum and symphysis to the transit time. Reference values of PWV were those estimated for Hispanic populations^[12]^.

#### Glucose and lipid profiles, hs CRP, and derived measurements

After 8–12 h overnight fast, total serum glucose, insulin, total cholesterol, triglycerides (TG), high-density lipoprotein cholesterol (HDL-chol), and high-sensitivity C-reactive protein (hs-CRP) were measured. Glucose (mg/dl) was measured with an enzymatic colorimetric test (QCA SA, Amposta, Spain), and radioimmunoassay (Diagnostic Products Corporation, Los Angeles, CA) was used for insulin determination (uUI/l). Dry analytical methodology determined the cholesterol profile (Vitros; Ortho Clinical Diagnostics Inc, Raritan, NJ). Serum hs-CRP (mg/l) was measured with a sensitive latex-based immunoassay, and values of >1.0 mg/l were considered low-grade systemic inflammation, according to the AHA/CDC Joint Statement on Markers of Inflammation and Cardiovascular Disease^[13]^. To avoid abnormally high levels of hs-CRP (denoting acute inflammation), participants being ill (e.g., cold, viral infections, diarrhea, etc.) at least ten days before the evaluation had their appointment rescheduled. The homeostatic model assessment (HOMA-IR) quantified insulin sensitivity, with values ≥2.6 denoting insulin resistance (IR)^[14]^. We also estimated HOMA-β and the Disposition Index to approach the functioning of the pancreatic β-cell. Metabolic Syndrome (MetS) was diagnosed based on the 2009 AHA/NHLBI/IDF Joint Interim Statement^[15]^. A continuous Metabolic Syndrome severity risk score was computed, according to Gurka et al., with values ≥ 1 SD considered to be of risk^[16]^. The Single-point Insulin Sensitivity Estimator was estimated using BMI, TG, and HDL, according to Paulmichl et al.^[17]^

### Imaging procedures for assessment of the liver and carotid arteries

#### Liver ultrasound

An abdominal ultrasound was performed on SLS participants using a General Electric LogiQ ultrasonographer with a 4C RS convex multifrequency probe (2–5.5 MHz) (GE Healthcare Systems, Wauwatosa, WI). All examinations were done by the same operator, who obtained and stored the images to be analyzed by two independent observers. The operator had training to obtain standardized images in which the liver and the right kidney were seen simultaneously. Images were obtained with the participant rolled onto its left side in a decubitus position, with the right arm stretched above the head after taking a deep breath. Observers were gastroenterologists with training in abdominal ultrasound interpretation and scored liver brightness (0-3), diaphragm attenuation (0-2), and vessel blurring (0-1), according to Hamaguchi et al^[18]^. The maximum score possible is six. According to the validity assessment of this semi-quantitative method to determine liver fat infiltration and conducted in the SLS with Magnetic Resonance Spectroscopy (MRS) as the standard, values ≥4 have good sensitivity (82%) and specificity (84%) for non-alcoholic fatty liver diseases diagnosis^[19]^.

#### Carotid ultrasound

Carotid intima-media thickness (CIMT) is an ultrasound-based technique that measures the intima and media thickness and examines the presence of carotid plaque. During the examination, each patient was lying down with their head tilted to the side (recumbent), and the neck fully exposed. A General Electric LogiQ ultra-sonographer with a 4C RS convex multifrequency probe (2–5.5 MHz) (GE Healthcare Systems, Wauwatosa, WI) was used to scan the neck. The probe was placed on the neck and moved upwards along the anterior/posterior edge of the sternocleidomastoid muscle. Two-dimensional images of the transverse and longitudinal axes were selected to observe the external and internal carotid arteries. The scan covered the transverse and longitudinal axes of the extracranial segment from the internal carotid artery to the common carotid artery. The examination determined the presence of plaque in the carotid artery tube, the location and size of the plaque, blood vessel stenosis, and blood flow. The technique and classification of CIMT follow the standards outlined in the consensus statement from the American Society of Echocardiography (ASE) Carotid Intima-Media Thickness Task Force^[20]^. Age, sex, and ethnicity were considered while determining the 75th percentile of risk, consistent with the ASE CIMT Task Force. If the IMT measurement is greater than the 75th percentile or if a carotid plaque is present, patients are at high risk of developing a cardiovascular event.

### Determination of age-related proteins

Blood samples were collected from the cohort and centrifuged to separate plasma, stored in 250 ul aliquots at -80°C. The Luminex system was used to determine the levels of the following proteins: IGF-1, IGF-2, OSM, myostatin, FGF-21, GDF15, GDF11, Apelin, Musclin, Sparc, Irisin, IL-2, IL-6, IL-10, and TNFα. To perform the Luminex assay, a Bioplex 200 platform (Bio-Rad Laboratories, California, USA) was used with the following kits: MILLIPLEX® MAP Human Cytokine-Chemokine Bead Panel (IL-2, IL-6, IL-10, and TNF-α), MILLIPLEX® Human Myokine Magnetic Bead Panel (Apelin, Myostatin, Irisin, Osteocrin, Osteonectin and Oncostatin), MILLIPLEX® Human Aging Magnetic Bead Panel 1 -Metabolism Multiplex Assay (GDF-11, GDF,15, FGF-21 and Leptin), and MILLIPLEX® Human IGF-I, II Magnetic Bead Panel (IGF-1, IFG-2) (Merck Millipore, Darmstadt, Germany). All reagents were applied and prepared following the manufacturer’s guidelines in each case. Namely, the plate was incubated with wash buffer for 10 min (shaking at RT). The washing buffer was then removed by vacuum filtration, and 25 μl of pre-coated mixed beads was added to each well and rinsed twice. Next, 50 μl of the prepared standard with a range of 0.13 pg/ml to 2000 pg/ml, together with the two provided kit controls, were added. Next, 50 μl of assay buffer was added to the background control well, and sample wells and 50 μl of the sample was added to the wells. The plate was then sealed and incubated at 4 °C (shaking) overnight. The unbound well content was removed the following day, and the beads were washed twice with a washing buffer. Next, 50 μl of detection antibodies solution was added and incubated for 1 h (RT). The beads were then incubated with 50 μl of Streptavidin-Phycoerythin solution for 30 min (RT). After two wash steps, 150 μl of sheath fluid (Bio-Rad) was added, and the signal was read with Bioplex manager software. All samples were measured in duplicate.

### Age-related epigenetic changes and telomeres length

Participants provided a 25 ml blood sample drawn into EDTA tubes during a morning assessment. The same morning, PBMCs were separated with a Ficoll-Paque density gradient (Ficoll-Paque™ Premium, cat No. 17-5442-03, GE Healthcare Bio-Sciences AB, Uppsala, Sweden). Next, DNA was extracted from PBMCs using the DNeasy Blood & Tissue Kit (Qiagen, Venlo, Netherlands), according to the manufacturer’s instructions. One µg of purified DNA was sent in batches of 96 to The Clock Foundation (Torrance, CA), preserving the cold chain. The Illumina Infinium MethylationEPIC BeadChip array was used. This array allows quantitatively examining over 850,000 CpG methylation locations per sample throughout the genome. Aside from computing well-established 1st and 2nd generation epigenetic aging clocks, it also provides a methylation-based estimation of leukocyte telomere length.

### Statistical analysis

Statistical analyses were conducted using Stata for Windows v.16.0 and XLSTAT-R, an interface between XLSTAT and the open-source R software. Data were expressed as mean (SD) or median (IQR) for descriptive purposes, depending on the normality of distribution. *Between-group comparison*: We used one-way ANOVA and Kruskal-Wallis H tests for statistical analysis. To estimate the impact of lifetime BMI trajectory on various health and aging markers, we calculated Cohen’s f and epsilon^2^, as effect size measures. *Within-group comparison*: Values of DNAmAge and chronological age of the same individual were compared by two-tailed paired Student’s *t*-test; Hedges *g* for paired data was computed as a measure of effect size. ANOVA for repeated measures allowed the comparison of anthropometric and cardiometabolic mean values of correlated samples. *Homogeneity in a data series*: We conducted the Pettit test for Change-Point Detection to examine whether the DNAmAge series was consistent across the entire sample. This analysis aimed to identify any possible change points within the series that could be attributed to the life-course BMI trajectory. By assessing the homogeneity of the series, we can gain a better understanding of the fundamental patterns and trends of the data. *Hierarchical cluster analysis*: Hierarchical clustering was conducted on standardized cardiometabolic and aging-related data (z score). Distance between clusters was computed based on the length of the straight line drawn from one cluster to another (Euclidean distance). We relied on Ward’s method as linkage criteria, which creates groups based on reducing the sum of squared distances of each observation from the average observation in a cluster. Heatmaps, dendrograms and hierarchical edge bundles were presented for males and females. Adjacency matrices are also provided. Of note, for the purpose of analysis, the values of HDL cholesterol, DNAm TL, IGF-1, IGF-2, Musclin, and CD4|CD8 ratio were the multiplicative inverse (1|x).

#### Predicting DNAm age from historic BMI data

Many prediction models only predict a single outcome measurement without considering the changes over time. However, by using repeated measurements of an independent variable, a model can gain more information about the variable’s development and trajectory over time. This information could be particularly useful in prediction modeling when the trajectory of an independent variable is more strongly associated with the outcome than a single measurement. By considering this trajectory, we may improve the accuracy of predicting this particular outcome. Based on the study by Welten et al.^[21]^, we developed a model to predict DNAmAge at the time of assessment using BMI measurements from birth to adulthood as a predictor. We conducted a two-step analysis based on standardized BMI (BAZ) estimated at various ages ranging from birth to last clinical assessment. Firstly, we performed linear regression with age as the independent variable and the BAZ measurement as the dependent variable. This regression analysis was conducted for each subject separately in a long-structured dataset to obtain individual growth curves from birth to adulthood over this period. We also applied quadratic and cubic functions besides a linear growth curve. Next, a new model was created by combining the mean of all BAZ measurements and the participant-specific growth parameters, i.e., slope coefficient(s), to predict the outcome DNAmAge at test time. Although Welten et al. used OLS-based estimators, we instead used multilevel models to better account for between-and within-group differences. In both cases, the mean value represents an individual’s average BAZ from the age of 0 to 28-29y. On the other hand, the slope parameters indicate the trend of an individual’s change in BAZ over time. An improved version of this model adjusted mean BAZ for the lifetime BMI trajectory group while including age at test time adjusted for sex. In all analyses, results were considered significant when a *p-value* of < 0.05 was obtained.

## DECLARATIONS

### Ethics approval

The IRBs of the Institute of Nutrition and Food Technology, University of Chile, obtained ethical approval (resolution date: December 1, 2021; number: 21–037). According to the norms for human experimentation, informed and written consent was obtained from all individual participants included in the study.

### Trial registration

The study does not consider any healthcare intervention on human participants.

### Consent for publication

Not applicable.

### Availability of data

The data supporting this study’s findings are available on request from the corresponding authors. The data are not publicly available due to privacy or ethical restrictions.

### Authors’ contribution

PC is the study’s lead investigator and obtained the necessary funds. RB and CGB also contribute to funding. PC, RB, CA, and CGB designed the study. PC, RB, FS, CGB conceptualized the fieldwork. CA, FS, and GS made significant contributions to the study protocol. RB coordinated fieldwork and management of clinical. RB and CS made substantial contributions to the obtention of clinical data. CGB, FS, and RT significantly contributed to lab data acquisition. RB and FS were responsible for interpreting clinical data. CGB is responsible for designing all laboratory methods of ObAGE. PC ran the data analysis, and CGB, FS, and RT assisted with interpreting the results. PC and CBG wrote the manuscript. All authors substantially revised this manuscript and approved the submitted version of the paper.

## REFERENCES

1. Khan, S., Ning, H., Wilkins, J, et al. Association of Body Mass Index with Lifetime Risk of Cardiovascular Disease and Compression of Morbidity. JAMA Cardiol 3, 280–287 (2018). DOI:10.1001/jamacardio.2018.0022

2. Iyen, B., Weng, S., Vinogradova, Y., Akyea, R., Qureshi, N. & Kai, J. Long-term body mass index changes in overweight and obese adults and the risk of heart failure, cardiovascular disease and mortality: a cohort study of over 260,000 adults in the UK. BMC Public Health 21, 576 (2021). DOI:10.1186/s12889-021-10606-1

3. Friedenreich, C., Ryder-Burbidge, C. & McNeil, J. Physical activity, obesity and sedentary behavior in cancer etiology: epidemiologic evidence and biologic mechanisms. Mol Oncol 15, 790–800 (2021). DOI:10.1002/1878-0261.12772

4. Cotangco, K., Liao, C., Eakin, C., et al. Trends in Incidence of Cancers Associated With Obesity and Other Modifiable Risk Factors Among Women, 2001-2018. Prev Chronic Dis 20, E21 (2023). DOI:10.5888/pcd20.220211

5. Chen, N., Fong, D. & Wong, J. Health and Economic Outcomes Associated With Musculoskeletal Disorders Attributable to High Body Mass Index in 192 Countries and Territories in 2019. JAMA Netw Open 6, e2250674 (2023). DOI:10.1001/jamanetworkopen.2022.50674

6. Cizza, G., Brown, R., Rother, K. Rising incidence and challenges of childhood diabetes. A mini review. J Endocrinol Invest 35, 541–546 (2012). DOI: 10.3275/8411.

7. Nadeau, K., Anderson, B., Berg, G., et al. Youth-Onset Type 2 Diabetes Consensus Report: Current Status, Challenges, and Priorities. Diabetes Care 39, 1635–1642 (2016). DOI: 10.2337/dc16-1066.

8. Kim, B., Kim, M., Han, K., et al. Low muscle mass is associated with metabolic syndrome only in nonobese young adults: the Korea National Health and Nutrition Examination Survey 2008-2010. Nutr Res 35, 1070–8 (2015). DOI: 10.1016/j.nutres.2015.09.020

9. Burrows, R., Correa, P., Reyes, M., Blanco, E., Albala, C. & Gahagan, S. Low muscle mass is associated with cardiometabolic risk regardless of nutritional status in adolescents: A cross-sectional study in a Chilean birth cohort. Pediatr Diabetes 18, 895–902 (2017). DOI: 10.1111/pedi.12505.

10. Marinac, C. & Birmann, B. Rising cancer incidence in younger adults: is obesity to blame? Lancet Public Health 4, e119–e120 (2019). DOI:10.1016/S2468-2667(19)30017-9

11. Salvestrini, V., Sell, C. & Lorenzini, A. Obesity May Accelerate the Aging Process. Front Endocrinol 3, 266 (2019). DOI: 10.3389/fendo.2019.00266.

12. Tam, B., Morais, J. & Santosa, S. Obesity and ageing: Two sides of the same coin. Obe Rev 21, e12991 (2020). DOI:10.1111/obr.12991

13. López-Otín, C., Blasco, M., Partridge., L., Serrano, M., Kroemer, G. Hallmarks of aging: An expanding universe. Cell 186, 243–278 (2023). DOI: 10.1016/j.cell.2022.11.001

14. Global BMI Mortality Collaboration. Body-mass index and all-cause mortality: individual-participant-data meta-analysis of 239 prospective studies in four continents. Lancet 388, 776–786 (2016). DOI: 10.1016/S0140-6736(16)30175-1.

15. Flegal, K., Kit, B., Orpana, H. & Graubard, B. Association of all-cause mortality with overweight and obesity using standard body mass index categories: a systematic review and meta-analysis. JAMA 309, 71–82 (2013). DOI: doi: 10.1001/jama.2012.113905

16. Berrington de Gonzalez, A., Hartge, P., Cerhan J., et al. Body-mass index and mortality among 1.46 million white adults. N Engl J Med 363, 2211–2219 (2010). DOI: 10.1056/NEJMoa1000367.

17. Reilly, J. & Kelly, J. Long-term impact of overweight and obesity in childhood and adolescence on morbidity and premature mortality in adulthood: systematic review. Int J Obes 35, 891–898 (2011). DOI: 10.1038/ijo.2010.222.

18. Fülöp, T., Larbi, A., Witkowski, J. Human Inflammaging. Gerontology 65, 495–504 (2019). doi: 10.1159/000497375.

19. Ley, R., Bäckhed, F., Turnbaugh, P., Lozupone, C., Knight, R. & Gordon, J. Obesity alters gut microbial ecology. Proc Natl Acad Sci 102, 11070–11075 (2005). DOI:10.1073/pnas.0504978102

20. Zhou, Y., Hambly, B. & McLachlan, C. FTO associations with obesity and telomere length. J Biomed Sci 24, 65 (2017). DOI: 10.1186/s12929-017-0372-6

21. De Mello, A., Costa, A., Engel, J., Rezin, G. Mitochondrial dysfunction in obesity. Life Sci 192, 26–32 (2018). DOI: 10.1016/j.lfs.2017.11.019.

22. Wen, X., Zhang, B., Wu, B. et al. Signaling pathways in obesity: mechanisms and therapeutic interventions. Signal Transduct Target Ther 7, 369 (2022). DOI: 10.1038/s41392-022-01188-4.

23. Bartelt, A. & Widenmaier, S. Proteostasis in thermogenesis and obesity. Biol Chem 401, 1019–1030 (2020). doi:10.1515/hsz-2019-0427

24. Newsholme, P. & de Bittencourt, P. The fat cell senescence hypothesis: a mechanism responsible for abrogating the resolution of inflammation in chronic disease. Curr Opin Clin Nutr Metab Care 17, 295–305. DOI: 10.1097/MCO.0000000000000077.

25. Franceschi, C. Healthy ageing in 2016: Obesity in geroscience -is cellular senescence the culprit? Nat Rev Endocrinol 13, 76–78 (2017). DOI: 10.1038/nrendo.2016.213.

26. Horvath, S., Erhart, W., Brosch M, et al. Obesity accelerates epigenetic aging of human liver. Proc Natl Acad Sci U S A 111, 15538–15543 (2014). DOI: 10.1073/pnas.1412759111.

27. De Toro-Martín, J., Guénard, F., Tchernof, A, et al. Body mass index is associated with epigenetic age acceleration in the visceral adipose tissue of subjects with severe obesity. Clin Epigenetics 11, 172 (2019). DOI: 10.1186/s13148-019-0754-6.

28. Lobstein T, Brindsen H. Obesity: missing the 2025 global targets -Trends, Costs and Country Reports. London: World Obesity Federation, 2020. Retrieved from: https://s3-eu-west-1.amazonaws.com/wof-files/970_-_WOF_Missing_the_2025_Global_Targets_Report_ART.pdf. Last accessed: June 2024.

29. Lozoff, B., De Andraca, I., Castillo, M., Smith, J., Walter, T. & Pino, P. Behavioral and developmental effects of preventing iron-deficiency anemia in healthy full-term infants. Pediatrics 112, 846–854 (2003). PMID: 14523176.

30. Correa, P., Rogan, J., Blanco, E., East, P., Lozoff, B., Gahagan, S. & Burrows R. Resolving early obesity leads to a cardiometabolic profile within normal ranges at 23 years old in a two-decade prospective follow-up study. Sci Rep 11, 18927 (2021). DOI: 10.1038/s41598-021-97683-9.

31. Lu, A., Seeboth, A., Tsai, P., et al. DNA methylation-based estimator of telomere length. Aging 11, 5895–5923 (2019). DOI: 10.18632/aging.102173.

32. Correa, P., Matamoros, M., de Toro, V., Zepeda, D., Arriaza, M. & Burrows, R. A Single-Point Insulin Sensitivity Estimator (SPISE) of 5.4 is a good predictor of both metabolic syndrome and insulin resistance in adolescents with obesity. Front Endocrinol 14, 1078949 (2023). DOI: 10.3389/fendo.2023.1078949.

33. Pedersen, B., Akerström, T., Nielsen, A. & Fischer C. Role of myokines in exercise and metabolism. J Appl Physiol 103, 1093–1098 (2007). DOI: 10.1152/japplphysiol.00080.2007.

34. Wang, B., Liang, J., Lu, C., Lu, A. & Wang, C. Exercise Regulates Myokines in Aging-Related Diseases through Muscle-Brain Crosstalk. Gerontology 70, 193–209 (2024). DOI: 10.1159/000535339.

35. López-Otín, C., Blasco, M., Partridge, L., Serrano, M. & Kroemer G. The hallmarks of aging. Cell 153, 1194–217 (2013). DOI: 10.1016/j.cell.2013.05.039.

36. González-Gualda, E., Baker, A., Fruk, L. & Muñoz-Espín, D. A guide to assessing cellular senescence in vitro and in vivo. FEBS J 288, 56–80 (2021). DOI: 10.1111/febs.15570.

37. Burtscher, J., Soltany, A., Visavadiya, N., Burtscher, M., Millet, G., Khoramipour, K. & Khamoui A. Mitochondrial stress and mitokines in aging. Aging Cell 22, e13770 (2023). DOI: 10.1111/acel.13770.

38. Welten, M., de Kroon, M., Renders, C., Steyerberg, E., Raat, H., Twisk, J., Heymans, M. Repeatedly measured predictors: a comparison of methods for prediction modeling. Diagn Progn Res. 2, 5 (2018). DOI: 10.1186/s41512-018-0024-7.

39. Nunan, E., Wright, C., Semola, O., et al. Obesity as a premature aging phenotype -implications for sarcopenic obesity. Geroscience 44, 1393–1405 (2022). DOI: 10.1007/s11357-022-00567-7

40. Oblak, L., van der Zaag, J., Higgins-Chen, A., Levine, M. & Boks, M. A systematic review of biological, social and environmental factors associated with epigenetic clock acceleration. Ageing Res Rev 69, 101348 (2021). DOI: 10.1016/j.arr.2021.101348.

41. Mundstock, E., Sarria, E., Zatti, H., et al. Effect of obesity on telomere length: Systematic review and meta-analysis. Obesity 23, 2165–2174 (2015). DOI: 10.1002/oby.21183.

42. Muhammad, T., Wan, Y., Sha, Q., et al. IGF2 improves the developmental competency and meiotic structure of oocytes from aged mice. Aging 13, 2118–2134 (2020). DOI: 10.18632/aging.202214.

43. Zhou, X., Tan, B., Gui, W., Zhou, C., Zhao, H., Lin, X. & Li, H. IGF2 deficiency promotes liver aging through mitochondrial dysfunction and upregulated CEBPB signaling in D-galactose-induced aging mice. Mol Med. 2023 Nov 28;29(1):161. DOI: 10.1186/s10020-023-00752-0.

44. Milman, S., Atzmon, G., Huffman, D., Wan, J., Crandall, J., Cohen, P. & Barzilai, N. Low insulin-like growth factor-1 level predicts survival in humans with exceptional longevity. Aging Cell. 13(4):769–771 (2014). DOI: 10.1111/acel.12213.

45. Zhang, W. & Milman, S. Looking at IGF-1 through the hourglass. Aging 14, 6379–6380 (2022). DOI: 10.18632/aging.204257.

46. Zhang, W., Ye, K., Barzilai, N. & Milman, S. The antagonistic pleiotropy of insulin-like growth factor 1. Aging Cell 20, e13443 (2021). DOI: 10.1111/acel.13443.

47. Gubbi, S., Quipildor, G., Barzilai, N., Huffman, D. & Milman, S. 40 YEARS of IGF1: IGF1: the Jekyll and Hyde of the aging brain. J Mol Endocrinol 61, T171–T185 (2018). DOI: 10.1530/JME-18-0093.

48. Añón-Hidalgo, J., Catalán, V., Rodríguez, A., et al. Circulating GDF11 levels are decreased with age but are unchanged with obesity and type 2 diabetes. Aging 11, 1733–1744 (2019). DOI: 10.18632/aging.101865.

49. Pedersen, L. & Hojman, P. Muscle-to-organ cross-talk mediated by myokines. Adipocyte 1, 64–167 (2012). DOI: 10.4161/adip.20344.

50. Severinsen, M. & Pedersen, B. Muscle-Organ Crosstalk: The Emerging Roles of Myokines. Endocr Rev 41, 594–609 (2020). DOI: 10.1210/endrev/bnaa016.

51. Jia, J., Yu, F., Wei, W., Yang, P., Zhang, R., Sheng, Y. & Shi, Y. Relationship between circulating irisin levels and overweight/obesity: A meta-analysis. World J Clin Cases 7, 1444–1455 (2019). DOI: 10.12998/wjcc.v7.i12.1444.

52. Li, C., Cheng, H., Adhikari, B., et al. The Role of Apelin-APJ System in Diabetes and Obesity. Front Endocrinol 13, 820002 (2022). DOI: 10.3389/fendo.2022.820002.

53. Yousefzadeh, M., Flores, R., Zhu, Y., et al. An aged immune system drives senescence and ageing of solid organs. Nature 94: 100–105 (2021). DOI: 10.1038/s41586-021-03547-7.

54. Burrows, R., Correa P., Rogan, J., Cheng, E., Blanco, E., Gahagan, S. Long-term *vs*. recent-onset obesity: their contribution to cardiometabolic risk in adolescence. Pediatr Res. 86, 776–782 (2019). DOI: 10.1038/s41390-019-0543-0.

55. Mattsson, M., Maher, G., Boland, F., Fitzgerald, A., Murray, D. & Biesma, R. Group-based trajectory modelling for BMI trajectories in childhood: A systematic review. Obes Rev 20, 998–1015 (2019). DOI: 10.1111/obr.12842.

56. Rabheru, K., Byles, J. & Kalache, A. How "old age" was withdrawn as a diagnosis from ICD-11. Lancet Healthy Longev 3, e457–e459 (2022). DOI: 10.1016/S2666-7568(22)00102-7.

57. López-Otín, C. & Kroemer, G. Hallmarks of Health. Cell 184, 33–63 (2021). DOI: 10.1016/j.cell.2020.11.034.

58. Cohen, A. Complex systems dynamics in aging: new evidence, continuing questions. Biogerontology 17, 205–220 (2016). DOI: 10.1007/s10522-015-9584-x.

59. Cohen A., Ferrucci, L., Fülöp, T., et al. A complex systems approach to aging biology. Nat Aging 2, 580–591. DOI: 10.1038/s43587-022-00252-6.

## References

1. World Medical Association. World Medical Association Declaration of Helsinki: ethical principles for medical research involving human subjects. JAMA 310, 2191–2194 (2013). DOI:10.1001/jama.2013.281053

2. Lozoff, B., De Andraca, I., Castillo, M., Smith, J., Walter, T. & Pino, P. Behavioral and developmental effects of preventing iron-deficiency anemia in healthy full-term infants. Pediatrics 112, 846–854 (2003). PMID: 14523176.

3. Albala, C., Vio, F., Kain, J. & Uauy, R. Nutrition transition in Chile: determinants and consequences. Public Health Nutr 5, 123–128 (2002). DOI: 10.1079/PHN2001283.

4. Albala, C., Vio, F., Kain, J. & Uauy, R. Nutrition transition in Latin America: the case of Chile. Nutr Rev 59, 170–176 (2001). DOI: 10.1111/j.1753-4887.2001.tb07008.x.

5. Sun, W., Joffe, M., Chen, J. & Brunelli, S. Design and analysis of multiple events case-control studies. Biometrics 66, 1220–1229 (2010). doi:10.1111/j.1541-0420.2009.01369.x

6. Correa, P., Burrows, R., Albala, C., et al. Multiple events case-control study in a prospective cohort to identify systemic, cellular, and molecular biomarkers of obesity-induced accelerated aging in 30-years-olds: the ObAGE study protocol. BMC Geriatr 22, 387 (2022). DOI: 10.1186/s12877-022-03032-4

7. De Onis, M. et al. Development of a WHO growth reference for school-aged children and adolescents. Bull. World Health Organ. 85, 660–667 (2007). DOI: 10.2471/blt.07.043497

8. Emery, W. & Thompson, R. Statistical Methods and Error Handling. in: Data Analysis Methods in Physical Oceanography. 193–304 (Elsevier, 2001). DOI: 10.1016/B978-044450756-03/50004-6.

9. Pollock, D. (1999). Smoothing with Cubic Splines. in: Handbook of Time Series Analysis, Signal Processing, and Dynamics. 293–322 (Academic Press, 1999). DOI: 10.1016/B978-012560990-6/50013-0.

10. Correa, P., Rogan, J., Blanco, E., East, P., Lozoff, B., Gahagan, S. & Burrows R. Resolving early obesity leads to a cardiometabolic profile within normal ranges at 23 years old in a two-decade prospective follow-up study. Sci Rep 11, 18927 (2021). DOI: 10.1038/s41598-021-97683-9.

11. Whelton, P., Carey, R., Aronow, F., et al. Guideline for the prevention, detection, evaluation, and management of high blood pressure in adults: a report of the American College of Cardiology/American Heart Association Task Force on Clinical Practice Guidelines Hypertension 71, e13–e115 (2018). DOI: 10.1161/HYP.0000000000000065

12. Bia, D. & Zócalo, Y. Physiological Age-and Sex-Related Profiles for Local (Aortic) and Regional (Carotid-Femoral, Carotid-Radial) Pulse Wave Velocity and Center-to-Periphery Stiffness Gradient, with and without Blood Pressure Adjustments: Reference Intervals and Agreement between Methods in Healthy Subjects (3-84 Years). J Cardiovasc Dev Dis 3 (2021). DOI:10.3390/jcdd8010003

13. Roberts WL; CDC; AHA. CDC/AHA Workshop on Markers of Inflammation and Cardiovascular Disease: Application to Clinical and Public Health Practice: A background paper: laboratory tests available to assess inflammation--performance and standardization. Circulation 110, e572–e576 (2004). DOI:10.1161/01.CIR.0000148986.52696.07

14. Burrows, R., Correa, P., Reyes. M., Blanco, E., Albala, C., Gahagan, S. Healthy Chilean Adolescents with HOMA-IR ≥ 2.6 Have Increased Cardiometabolic Risk: Association with Genetic, Biological, and Environmental Factors. J Diabetes Res 2015, 783296 (2015). DOI:10.1155/2015/783296

15. Alberti KG, Eckel RH, Grundy SM, et al. Harmonizing the metabolic syndrome: a joint interim statement of the International Diabetes Federation Task Force on Epidemiology and Prevention; National Heart, Lung, and Blood Institute; American Heart Association; World Heart Federation; International Atherosclerosis Society; and International Association for the Study of Obesity. Circulation. 120, 1640–1645 (2009). DOI:10.1161/CIRCULATIONAHA.109.192644

16. DeBoer, M. & Gurka, M. Clinical utility of metabolic syndrome severity scores: considerations for practitioners. Diabetes Metab Syndr Obes 10, 65–72 (2017). DOI: 10.2147/DMSO.S101624.

17. Paulmichl, K., Hatunic, M., Højlund, K., et al. Modification and Validation of the Triglyceride-to-HDL Cholesterol Ratio as a Surrogate of Insulin Sensitivity in White Juveniles and Adults without Diabetes Mellitus: The Single Point Insulin Sensitivity Estimator (SPISE). Clin Chem 62, 1211–1219 (2016). DOI: 10.1373/clinchem.2016.257436

18. Hamaguchi, M., Kojima, T., Itoh, Y., et al. The severity of ultrasonographic findings in nonalcoholic fatty liver disease reflects the metabolic syndrome and visceral fat accumulation. Am J Gastroenterol 102, 2708–15 (2007). doi: 10.1111/j.1572-0241.2007.01526.x.

19. Ibacahe, C., Correa, P., Burrows, R., et al. Accuracy of a Semi-Quantitative Ultrasound Method to Determine Liver Fat Infiltration in Early Adulthood. Diagnostics 10, 431 (2020). DOI: 10.3390/diagnostics10060431.

20. Stein, J., Korcarz, C., Hurst, R., et al. Use of carotid ultrasound to identify subclinical vascular disease and evaluate cardiovascular disease risk: a consensus statement from the American Society of Echocardiography Carotid Intima-Media Thickness Task Force. J Am Soc Echocardiogr 21, 93–190 (2008). DOI:10.1016/j.echo.2007.11.011.

21. Welten, M., de Kroon, M., Renders, C., Steyerberg, E., Raat, H., Twisk, J., Heymans, M. Repeatedly measured predictors: a comparison of methods for prediction modeling. Diagn Progn Res. 2, 5 (2018). DOI: 10.1186/s41512-018-0024-7.

