## Supplemental Data for "Persistent obesity since childhood or adolescence accelerates biological aging in young adults from Chile’s oldest birth cohort"

**Table S1 Sample description by sex at 28y-assessment (n=205)**

| | Male participants (n=105) | | | Female participants (n=100) | | |
|---|---|---|---|---|---|---|
| | Mean \| Median | SD \| IQR | Range | Mean \| Median | SD \| IQR | Range |
| Chronological age (y) | 28.9 | 0.8 | 28.0 -31.3 | 28.9 | 0.6 | 28.0 – 31.0 |
| DNAm age (y) | 32.1 | 4.5 | 23.2 – 43.7 | 30.6 | 4.3 | 23.0 – 41.4 |
| Grim age (y) | 30.1 | 4.7 | 20.8 – 41.1 | 29.0 | 4.4 | 19.4 – 40.3 |
| Body-mass Index | 31.5 | 7.2 | 19.4 – 51.1 | 32.3 | 8.6 | 18.5 – 51.5 |
| Total body fat mass (%) | 34.8 | 7.7 | 13.6 – 50.0 | 43.2 | 8.1 | 21.5 – 56.2 |
| Total body lean mass (%) | 61.9 | 7.3 | 47.9 – 82.5 | 54.0 | 7.8 | 41.4 – 74.4 |
| Truncal fat mass (%) | 30.9 | 6.7 | 10.4 – 42.9 | 32.0 | 7.3 | 16.4 – 45.9 |
| Systolic BP (mm Hg) | *125* | *14* | 101 - 151 | *117* | *16* | 93 - 149 |
| Diastolic BP (mm Hg) | *77* | *12* | 59 - 98 | *73* | *13* | 53 - 109 |
| VOP (m/s) | 5.4 | 0.3 | 4.1 – 6.4 | 5.2 | 0.3 | 4.4 – 6.2 |
| CIMT left (mm) | 0.51 | 0.07 | 0.32 – 0.72 | 0.48 | 0.06 | 0.37 – 0.70 |
| CIMT right (mm) | 0.48 | 0.06 | 0.33 – 0.69 | 0.45 | 0.05 | 0.34 – 0.59 |
| Fasting glycemia (mg/dl) | 94.2 | 8.9 | 75.0 – 121.5 | 89.6 | 9.3 | 72.7 – 122.4 |
| Fasting insulin (uUI/L) | *12.8* | *8.4* | 4.7 – 24.5 | *11.3* | *7.6* | 5.0 – 23.7 |
| HOMA-IR | *2.75* | *2.1* | 1.1 – 8.0 | *2.55* | *2.45* | 1.0 – 9.3 |
| HOMA-β (%) | *137.4* | *88.2* | 54.4 – 298.4 | *164.8* | *112.8* | 34.2 – 297.5 |
| Hamagushi liver score (0-6) | *4* | *3* | 0 - 6 | *3* | *2* | 0 - 6 |
| **BMI category group** | | | | | | |
| TG1 | 46 | 43.3% | (…) | 43 | 43.0% | (…) |
| TG2 | 19 | 18.1% | (…) | 24 | 24.0% | (…) |
| TG3 | 40 | 38.1% | (…) | 33 | 33.0% | (…) |
| **Cardiometabolic-related alterations** | | | | | | |
| Insulin resistance | 61 | 58.1% | (…) | 57 | 57.0% | (…) |
| Metabolic syndrome | 74 | 70.5% | (…) | 40 | 40.0% | (…) |
| 3NAFLD | 54 | 51.9% | (…) | 42 | 42.4% | (…) |
| Low-grade systemic inflammation | 44 | 41.9% | (…) | 42 | 42.4% | (…) |
| Type-2 diabetes | 1 | 0.95% | (…) | 0 | 0.00% | (…) |
| Metformin | 1 | 0.95% | (…) | 3 | 3.00% | (…) |

Values are expressed as mean and SD, *median and IQR* or n and percentage.

**Table S2** Anthropometric phenotyping of ObAGE participants by BMI trajectory (n=205)

| | TG1 (n=89) | TG2 (n=43) | TG3 (n=73) | Between-group Differences* | Cohen's f |
|---|---|---|---|---|---|
| Chronological age (y) | 29.1 ± 0.4 | 28.7 ± 0.4 | 28.8 ± 0.4 | NS | … |
| Body-mass Index (kg/m2) | 23.1 ± 3.6 | 34.3 ± 4.4 | 37.7 ± 6.0 | ABC | 1.25 (1.05-1.35) |
| Waist circumference (cm) | 83.4 ± 10.7 | 100 ± 10.9 | 108.9 ± 13.9 | ABC | 0.95 (0.81-1.09) |
| Whole body fat mass (%) | 33.3 ± 7.2 | 43.0 ± 7.0 | 42.6 ± 7.7 | ABB | 0.62 (0.49-0.74) |
| Whole body lean mass (%) | 63.0 ± 6.8 | 54.0 ± 6.6 | 54.5 ± 7.2 | ABB | 0.60 (0.48-0.73) |
| Truncal fat mass (%) | 26.9 ± 5.9 | 34.6 ± 3.8 | 35.4 ± 5.1 | ABB | 0.75 (0.62-0.88) |
| Truncal fat mass \| Whole body fat mass | 0.50 ± 0.05 | 0.55 ± 0.04 | 0.57 ± 0.05 | ABB | 0.62 (0.49-0.77) |
| Legs FM \| Whole body fat mass | 0.35 ± 0.04 | 0.31 ± 0.04 | 0.29 ± 0.05 | ABB | 0.49 (0.36-0.63) |
| Appendicular fat mass \| Truncal fat mass | 0.91 ± 0.21 | 0.75 ± 0.14 | 0.69 ± 0.16 | ABB | 0.52 (0.39-0.66) |
| Lean Mass \| Fat Mass | 2.24 ± 0.81 | 1.25 ± 0.33 | 1.34 ± 0.48 | ABB | 0.73 (0.59-0.88) |
| Appendicular lean mass \| Append fat mass | 2.28 ± 0.93 | 1.45 ± 0.49 | 1.66 ± 0.66 | ABB | 0.45 (0.32-0.59) |

Values expressed as mean ± SD, *HSD Tukey post-hoc test: A=TG1; B=TG2; C=TG3. Different letters indicate significant statistical differences. The same letter denotes means that do not differ. The effect size for the difference was computed as Cohen's *f* coefficient: 0.10 = weak effect size, 0.25 = moderate effect size, and 0.40 = strong effect size. TG1 refers to participants always having a BMI in the healthy range; TG2 refers to participants with obesity starting in adolescence and remaining obese into adulthood; and TG3 refers to participants who were obese in early childhood and remained obese into adulthood (TG3).

**Table S3** Cardiometabolic phenotyping of ObAGE participants by BMI trajectory (n=205)

| | TG1 (n=89) | TG2 (n=43) | TG3 (n=73) | Between-group Differences* | $\varepsilon^2$** |
|---|---|---|---|---|---|
| Systolic Blood Pressure (mmHg) | 118 (19) | 122 (12) | 124 (16) | ABB | 0.05 (W) |
| Diastolic Blood Pressure (mmHg) | 75 (13) | 77 (12) | 76 (13) | NS | (…) |
| Pulse-wave velocity (m/s) | 5.1 ± 0.3 | 5.4 ± 0.3 | 5.4 ± 0.3 | ABB† | 0.09 (M)‡ |
| CIMT (left) (mm) | 0.48 ± .06 | 0.49 ± .07 | 0.50 ± .07 | NS† | (…) |
| CIMT (right) (mm) | 0.45 ± .06 | 0.47 ± .05 | 0.48 ± .07 | NS† | (…) |
| Fasting Glycemia (mg/dl) | 91.1 ± 7.6 | 92.1 ± 11.1 | 97.8 ± 32.6 | AAC† | 0.16 (VW)‡ |
| Fasting Insulin (uUI/l) | 10.4 (6.3) | 14.3 (9.0) | 16.7 (12.5) | ABB | 0.14 (S) |
| HOMA-IR | 2.3 (1.4) | 3.2 (2.3) | 3.9 (3.3) | ABB | 0.12 (M) |
| HOMA-β (%) | 133.4 (98.3) | 191.0 (123.5) | 191.7 (155.0) | ABB | 0.11 (M) |
| Triglycerides (mg/dl) | 84.1 (86.4) | 115.6 (75.0) | 121.3 (99.9) | ABB | 0.02 (W) |
| HDL cholesterol (mg/dl) | 31.6 (7.9) | 22.0 (9.6) | 20.1 (9.9) | NS | (…) |
| Total cholesterol (mg/dl) | 167.4 (53.4) | 178.9 (51.1) | 173.9 (59.8) | NS | (…) |
| MetS severity score (SD) | 0.12 (1.0) | 0.72 (0.8) | 1.0 (1.3) | ABC | 0.13 (M) |
| Hamagushi liver score (0-6) | 2 (2) | 4 (3) | 4 (3) | ABB | 0.14 (S) |

Values are expressed as mean ± SD or median (IQR). The effect size for the Kruskal-Wallis test is computed as the $\varepsilon^2$ based on the H-statistic: $\varepsilon 2 [H] = (H - k + 1)/(n - k)$, where H is the value obtained in the Kruskal-Wallis test; k is the number of groups; n is the total number of observations. The ε2 estimate assumes values from 0 to 1; multiplied by 100, indicates the percentage of variance in the dependent variable explained by the independent variable. The interpretation values commonly in published literature are 0.01- < 0.06 (weak effect), 0.06 - < 0.14 (moderate effect), and >= 0.14 (strong effect). The ES for the ANOVA test was computed as Cohen's *f* coefficient: 0.10 = weak effect size, 0.25 = moderate effect size, and 0.40 = strong effect size. TG1: participants always having a BMI in the healthy range; TG2: participants with obesity starting in adolescence and remaining obese into adulthood; TG3: participants who were obese in early childhood and remaining obese into adulthood. CIMT: carotid intima-media thickness. HOMA: Homeostatic Model Assessment. HDL: High-density lipoprotein. A=TG1; B=TG2; C=TG3. In both, ANOVA and K-W post hoc tests, different letters indicate significant statistical differences. The same letter denotes means or median do not differ.

**Fig. S1** Hierarchical clustering of anthropometric and cardiometabolic markers in males: phylogenetic dendrogram (n=105)

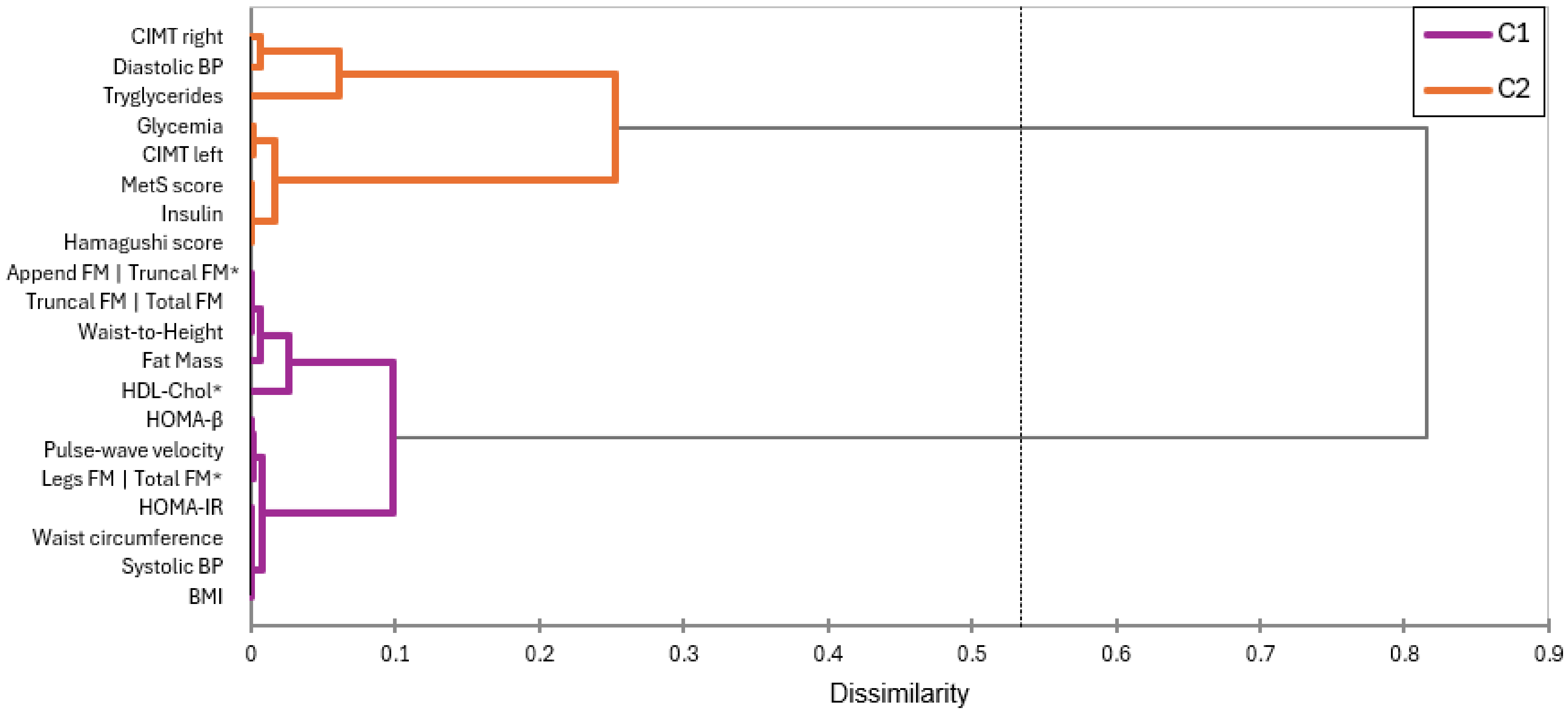


Hierarchical cluster analysis on standardized data of selected anthropometric and cardiometabolic markers assessed at 28- 29y. Distance criteria: Euclidian method. Linkage criteria: Ward linkage. BP: blood pressure. HDL: high-density lipoprotein. HOMA-IR: Homeostatic model assessment – Insulin Resistance. HOMA-β: Homeostatic model assessment – β cell function. CIMT: carotid intima-media thickness. FM: whole-body fat mass. HDL-chol, legs FM | total FM and appendicular FM | truncal FM are the multiplicative inverse (1/x). C1 refers to Cluster 1 and C2 refers to Cluster 2

**Table S4** Hierarchical clustering of anthropometric and cardiometabolic markers in males (n=105): adjacency matrix

<table>
<tr><th></th><th>BMI</th><th>Waist circumference</th><th>Waist-to-Height</th><th>Fat Mass</th><th>Truncal FM | Total FM</th><th>Legs FM | Total FM*</th><th>Appendicular FM | Truncal FM*</th><th>Systolic BP</th><th>Diastolic BP</th><th>Pulse-wave velocity</th><th>CIMT left</th><th>CIMT right</th><th>Glycemia</th><th>Insulin</th><th>HOMA-IR</th><th>HOMA-β</th><th>Triglycerides</th><th>HDL-Chol*</th><th>MetS score</th><th>Hamagushi score</th></tr>
<tr><td>BMI</td><td>0</td><td>0.005</td><td>0.014</td><td>0.046</td><td>0.022</td><td>0.011</td><td>0.019</td><td>0.005</td><td>0.278</td><td>0.006</td><td>0.068</td><td>0.173</td><td>0.093</td><td>0.031</td><td>0.006</td><td>0.007</td><td>0.577</td><td>0.103</td><td>0.033</td><td>0.027</td></tr>
<tr><td>Waist circumference</td><td>0.005</td><td>0</td><td>0.021</td><td>0.045</td><td>0.027</td><td>0.028</td><td>0.024</td><td>0.017</td><td>0.308</td><td>0.020</td><td>0.094</td><td>0.202</td><td>0.120</td><td>0.053</td><td>0.017</td><td>0.022</td><td>0.606</td><td>0.093</td><td>0.055</td><td>0.048</td></tr>
<tr><td>Waist-to-Height</td><td>0.014</td><td>0.021</td><td>0</td><td>0.011</td><td>0.001</td><td>0.031</td><td>0.000</td><td>0.007</td><td>0.398</td><td>0.015</td><td>0.127</td><td>0.267</td><td>0.161</td><td>0.070</td><td>0.004</td><td>0.020</td><td>0.753</td><td>0.049</td><td>0.073</td><td>0.063</td></tr>
<tr><td>Fat Mass</td><td>0.046</td><td>0.045</td><td>0.011</td><td>0</td><td>0.005</td><td>0.080</td><td>0.007</td><td>0.035</td><td>0.540</td><td>0.052</td><td>0.214</td><td>0.386</td><td>0.258</td><td>0.138</td><td>0.029</td><td>0.062</td><td>0.939</td><td>0.013</td><td>0.142</td><td>0.127</td></tr>
<tr><td>Truncal FM | Total FM</td><td>0.022</td><td>0.027</td><td>0.001</td><td>0.005</td><td>0</td><td>0.044</td><td>0.000</td><td>0.013</td><td>0.440</td><td>0.024</td><td>0.152</td><td>0.302</td><td>0.189</td><td>0.089</td><td>0.009</td><td>0.031</td><td>0.809</td><td>0.035</td><td>0.092</td><td>0.081</td></tr>
<tr><td>Legs FM | Total FM*</td><td>0.011</td><td>0.028</td><td>0.031</td><td>0.080</td><td>0.044</td><td>0</td><td>0.039</td><td>0.009</td><td>0.209</td><td>0.003</td><td>0.033</td><td>0.117</td><td>0.051</td><td>0.008</td><td>0.013</td><td>0.001</td><td>0.487</td><td>0.157</td><td>0.009</td><td>0.005</td></tr>
<tr><td>Appendicular FM | Truncal FM*</td><td>0.019</td><td>0.024</td><td>0.000</td><td>0.007</td><td>0.000</td><td>0.039</td><td>0</td><td>0.010</td><td>0.424</td><td>0.021</td><td>0.142</td><td>0.289</td><td>0.179</td><td>0.082</td><td>0.007</td><td>0.027</td><td>0.788</td><td>0.040</td><td>0.085</td><td>0.074</td></tr>
<tr><td>Systolic BP</td><td>0.005</td><td>0.017</td><td>0.007</td><td>0.035</td><td>0.013</td><td>0.009</td><td>0.010</td><td>0</td><td>0.304</td><td>0.002</td><td>0.076</td><td>0.191</td><td>0.104</td><td>0.034</td><td>0.000</td><td>0.004</td><td>0.625</td><td>0.091</td><td>0.036</td><td>0.029</td></tr>
<tr><td>Diastolic BP</td><td>0.278</td><td>0.308</td><td>0.398</td><td>0.540</td><td>0.440</td><td>0.209</td><td>0.424</td><td>0.304</td><td>0</td><td>0.261</td><td>0.078</td><td>0.014</td><td>0.054</td><td>0.137</td><td>0.323</td><td>0.242</td><td>0.060</td><td>0.712</td><td>0.132</td><td>0.148</td></tr>
<tr><td>Pulse-wave velocity</td><td>0.006</td><td>0.020</td><td>0.015</td><td>0.052</td><td>0.024</td><td>0.003</td><td>0.021</td><td>0.002</td><td>0.261</td><td>0</td><td>0.055</td><td>0.157</td><td>0.079</td><td>0.020</td><td>0.003</td><td>0.000</td><td>0.563</td><td>0.117</td><td>0.022</td><td>0.017</td></tr>
<tr><td>CIMT left</td><td>0.068</td><td>0.094</td><td>0.127</td><td>0.214</td><td>0.152</td><td>0.033</td><td>0.142</td><td>0.076</td><td>0.078</td><td>0.055</td><td>0</td><td>0.026</td><td>0.002</td><td>0.008</td><td>0.086</td><td>0.046</td><td>0.272</td><td>0.329</td><td>0.007</td><td>0.011</td></tr>
<tr><td>CIMT right</td><td>0.173</td><td>0.202</td><td>0.267</td><td>0.386</td><td>0.302</td><td>0.117</td><td>0.289</td><td>0.191</td><td>0.014</td><td>0.157</td><td>0.026</td><td>0</td><td>0.014</td><td>0.065</td><td>0.206</td><td>0.142</td><td>0.130</td><td>0.536</td><td>0.062</td><td>0.072</td></tr>
<tr><td>Glycemia</td><td>0.093</td><td>0.120</td><td>0.161</td><td>0.258</td><td>0.189</td><td>0.051</td><td>0.179</td><td>0.104</td><td>0.054</td><td>0.079</td><td>0.002</td><td>0.014</td><td>0</td><td>0.019</td><td>0.115</td><td>0.068</td><td>0.227</td><td>0.383</td><td>0.017</td><td>0.023</td></tr>
<tr><td>Insulin</td><td>0.031</td><td>0.053</td><td>0.070</td><td>0.138</td><td>0.089</td><td>0.008</td><td>0.082</td><td>0.034</td><td>0.137</td><td>0.020</td><td>0.008</td><td>0.065</td><td>0.019</td><td>0</td><td>0.041</td><td>0.015</td><td>0.374</td><td>0.234</td><td>0.000</td><td>0.000</td></tr>
<tr><td>HOMA-IR</td><td>0.006</td><td>0.017</td><td>0.004</td><td>0.029</td><td>0.009</td><td>0.013</td><td>0.007</td><td>0.000</td><td>0.323</td><td>0.003</td><td>0.086</td><td>0.206</td><td>0.115</td><td>0.041</td><td>0</td><td>0.006</td><td>0.651</td><td>0.081</td><td>0.043</td><td>0.035</td></tr>
<tr><td>HOMA-β</td><td>0.007</td><td>0.022</td><td>0.020</td><td>0.062</td><td>0.031</td><td>0.001</td><td>0.027</td><td>0.004</td><td>0.242</td><td>0.000</td><td>0.046</td><td>0.142</td><td>0.068</td><td>0.015</td><td>0.006</td><td>0</td><td>0.535</td><td>0.131</td><td>0.017</td><td>0.012</td></tr>
<tr><td>Triglycerides</td><td>0.577</td><td>0.606</td><td>0.753</td><td>0.939</td><td>0.809</td><td>0.487</td><td>0.788</td><td>0.625</td><td>0.060</td><td>0.563</td><td>0.272</td><td>0.130</td><td>0.227</td><td>0.374</td><td>0.651</td><td>0.535</td><td>0</td><td>1.155</td><td>0.367</td><td>0.392</td></tr>
<tr><td>HDL-Chol*</td><td>0.103</td><td>0.093</td><td>0.049</td><td>0.013</td><td>0.035</td><td>0.157</td><td>0.040</td><td>0.091</td><td>0.712</td><td>0.117</td><td>0.329</td><td>0.536</td><td>0.383</td><td>0.234</td><td>0.081</td><td>0.131</td><td>1.155</td><td>0</td><td>0.239</td><td>0.220</td></tr>
<tr><td>MetS score</td><td>0.033</td><td>0.055</td><td>0.073</td><td>0.142</td><td>0.092</td><td>0.009</td><td>0.085</td><td>0.036</td><td>0.132</td><td>0.022</td><td>0.007</td><td>0.062</td><td>0.017</td><td>0.000</td><td>0.043</td><td>0.017</td><td>0.367</td><td>0.239</td><td>0</td><td>0.000</td></tr>
<tr><td>Hamagushi score</td><td>0.027</td><td>0.048</td><td>0.063</td><td>0.127</td><td>0.081</td><td>0.005</td><td>0.074</td><td>0.029</td><td>0.148</td><td>0.017</td><td>0.011</td><td>0.072</td><td>0.023</td><td>0.000</td><td>0.035</td><td>0.012</td><td>0.392</td><td>0.220</td><td>0.000</td><td>0</td></tr>
</table>

Hierarchical cluster analysis on standardized data of selected anthropometric and cardiometabolic markers assessed at 28- 29y. Distance criteria: Euclidian method. Linkage criteria: Ward linkage. BP: blood pressure. HDL: high-density lipoprotein. HOMA-IR: Homeostatic model assessment – Insulin Resistance. HOMA-β: Homeostatic model assessment – β cell function. CIMT: carotid intima-media thickness. FM: whole-body fat mass. HDL-chol, legs FM | total FM and appendicular FM | truncal FM are the multiplicative inverse (1/x).

**Fig. S2** Hierarchical clustering of anthropometric and cardiometabolic markers in females (n=100): phylogenetic dendrogram

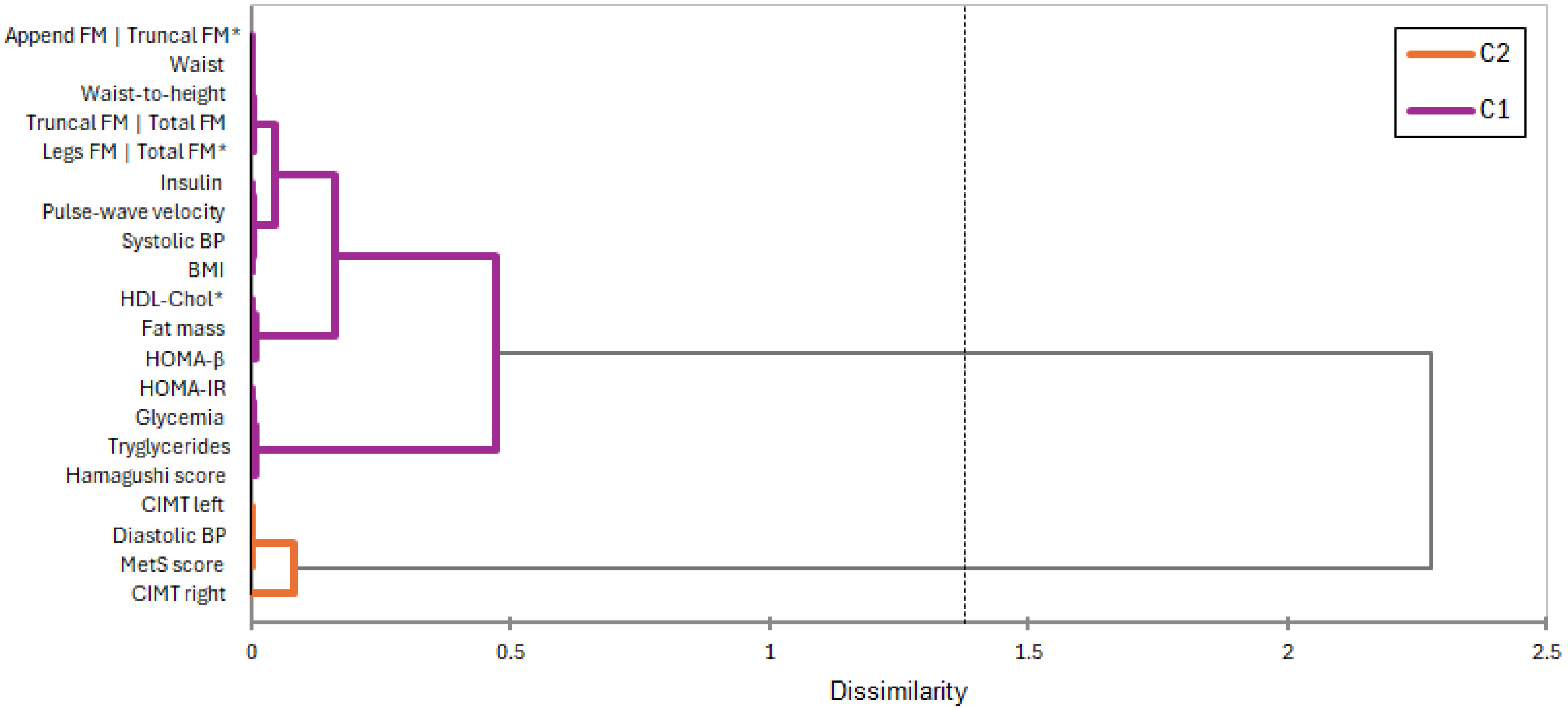


Hierarchical cluster analysis on standardized data of selected anthropometric and cardiometabolic markers assessed at 28- 29y. Distance criteria: Euclidian method. Linkage criteria: Ward linkage. BP: blood pressure. HDL: high-density lipoprotein. HOMA-IR: Homeostatic model assessment – Insulin Resistance. HOMA-β: Homeostatic model assessment – β cell function. CIMT: carotid intima-media thickness. FM: whole-body fat mass. HDL-chol, legs FM | total FM and appendicular FM | truncal FM are the multiplicative inverse (1/x). C1 refers to Cluster 1 and C2 refers to Cluster 2

### Table S5 Hierarchical clustering of anthropometric and cardiometabolic markers in females (n=100): adjacency matrix

| | BMI | Waist circumference | Waist-to-Height | Fat Mass | Truncal FM \| Total FM | Legs FM \| Total FM* | Appendicular FM \| Truncal FM* | Systolic BP | Diastolic BP | Pulse-wave velocity | CIMT left | CIMT right | Glycemia | Insulin | HOMA-IR | HOMA-β | Triglycerides | HDL-Chol* | MetS score | Hamagushi score |
|---|---|---|---|---|---|---|---|---|---|---|---|---|---|---|---|---|---|---|---|---|
| BMI | **0** | 0.005 | 0.014 | 0.046 | 0.022 | 0.011 | 0.019 | 0.005 | 0.278 | 0.006 | 0.068 | 0.173 | 0.093 | 0.031 | 0.006 | 0.007 | 0.577 | 0.103 | 0.033 | 0.027 |
| Waist circumference | 0.005 | **0** | 0.021 | 0.045 | 0.027 | 0.028 | 0.024 | 0.017 | 0.308 | 0.020 | 0.094 | 0.202 | 0.120 | 0.053 | 0.017 | 0.022 | 0.606 | 0.093 | 0.055 | 0.048 |
| Waist-to-Height | 0.014 | 0.021 | **0** | 0.011 | 0.001 | 0.031 | 0.000 | 0.007 | 0.398 | 0.015 | 0.127 | 0.267 | 0.161 | 0.070 | 0.004 | 0.020 | 0.753 | 0.049 | 0.073 | 0.063 |
| Fat Mass | 0.046 | 0.045 | 0.011 | **0** | 0.005 | 0.080 | 0.007 | 0.035 | 0.540 | 0.052 | 0.214 | 0.386 | 0.258 | 0.138 | 0.029 | 0.062 | 0.939 | 0.013 | 0.142 | 0.127 |
| Truncal FM \| Total FM | 0.022 | 0.027 | 0.001 | 0.005 | **0** | 0.044 | 0.000 | 0.013 | 0.440 | 0.024 | 0.152 | 0.302 | 0.189 | 0.089 | 0.009 | 0.031 | 0.809 | 0.035 | 0.092 | 0.081 |
| Legs FM \| Total FM* | 0.011 | 0.028 | 0.031 | 0.080 | 0.044 | **0** | 0.039 | 0.009 | 0.209 | 0.003 | 0.033 | 0.117 | 0.051 | 0.008 | 0.013 | 0.001 | 0.487 | 0.157 | 0.009 | 0.005 |
| Appendicular FM \| Truncal FM* | 0.019 | 0.024 | 0.000 | 0.007 | 0.000 | 0.039 | **0** | 0.010 | 0.424 | 0.021 | 0.142 | 0.289 | 0.179 | 0.082 | 0.007 | 0.027 | 0.788 | 0.040 | 0.085 | 0.074 |
| Systolic BP | 0.005 | 0.017 | 0.007 | 0.035 | 0.013 | 0.009 | 0.010 | **0** | 0.304 | 0.002 | 0.076 | 0.191 | 0.104 | 0.034 | 0.000 | 0.004 | 0.625 | 0.091 | 0.036 | 0.029 |
| Diastolic BP | 0.278 | 0.308 | 0.398 | 0.540 | 0.440 | 0.209 | 0.424 | 0.304 | **0** | 0.261 | 0.078 | 0.014 | 0.054 | 0.137 | 0.323 | 0.242 | 0.060 | 0.712 | 0.132 | 0.148 |
| Pulse-wave velocity | 0.006 | 0.020 | 0.015 | 0.052 | 0.024 | 0.003 | 0.021 | 0.002 | 0.261 | **0** | 0.055 | 0.157 | 0.079 | 0.020 | 0.003 | 0.000 | 0.563 | 0.117 | 0.022 | 0.017 |
| CIMT left | 0.068 | 0.094 | 0.127 | 0.214 | 0.152 | 0.033 | 0.142 | 0.076 | 0.078 | 0.055 | **0** | 0.026 | 0.002 | 0.008 | 0.086 | 0.046 | 0.272 | 0.329 | 0.007 | 0.011 |
| CIMT right | 0.173 | 0.202 | 0.267 | 0.386 | 0.302 | 0.117 | 0.289 | 0.191 | 0.014 | 0.157 | 0.026 | **0** | 0.014 | 0.065 | 0.206 | 0.142 | 0.130 | 0.536 | 0.062 | 0.072 |
| Glycemia | 0.093 | 0.120 | 0.161 | 0.258 | 0.189 | 0.051 | 0.179 | 0.104 | 0.054 | 0.079 | 0.002 | 0.014 | **0** | 0.019 | 0.115 | 0.068 | 0.227 | 0.383 | 0.017 | 0.023 |
| Insulin | 0.031 | 0.053 | 0.070 | 0.138 | 0.089 | 0.008 | 0.082 | 0.034 | 0.137 | 0.020 | 0.008 | 0.065 | 0.019 | **0** | 0.041 | 0.015 | 0.374 | 0.234 | 0.000 | 0.000 |
| HOMA-IR | 0.006 | 0.017 | 0.004 | 0.029 | 0.009 | 0.013 | 0.007 | 0.000 | 0.323 | 0.003 | 0.086 | 0.206 | 0.115 | 0.041 | **0** | 0.006 | 0.651 | 0.081 | 0.043 | 0.035 |
| HOMA-β | 0.007 | 0.022 | 0.020 | 0.062 | 0.031 | 0.001 | 0.027 | 0.004 | 0.242 | 0.000 | 0.046 | 0.142 | 0.068 | 0.015 | 0.006 | **0** | 0.535 | 0.131 | 0.017 | 0.012 |
| Triglycerides | 0.577 | 0.606 | 0.753 | 0.939 | 0.809 | 0.487 | 0.788 | 0.625 | 0.060 | 0.563 | 0.272 | 0.130 | 0.227 | 0.374 | 0.651 | 0.535 | **0** | 1.155 | 0.367 | 0.392 |
| HDL-Chol* | 0.103 | 0.093 | 0.049 | 0.013 | 0.035 | 0.157 | 0.040 | 0.091 | 0.712 | 0.117 | 0.329 | 0.536 | 0.383 | 0.234 | 0.081 | 0.131 | 1.155 | **0** | 0.239 | 0.220 |
| MetS score | 0.033 | 0.055 | 0.073 | 0.142 | 0.092 | 0.009 | 0.085 | 0.036 | 0.132 | 0.022 | 0.007 | 0.062 | 0.017 | 0.000 | 0.043 | 0.017 | 0.367 | 0.239 | **0** | 0.000 |
| Hamagushi score | 0.027 | 0.048 | 0.063 | 0.127 | 0.081 | 0.005 | 0.074 | 0.029 | 0.148 | 0.017 | 0.011 | 0.072 | 0.023 | 0.000 | 0.035 | 0.012 | 0.392 | 0.220 | 0.000 | **0** |

Hierarchical cluster analysis on standardized data of selected anthropometric and cardiometabolic markers assessed at 28- 29y. Distance criteria: Euclidian method. Linkage criteria: Ward linkage. BP: blood pressure. HDL: high-density lipoprotein. HOMA-IR: Homeostatic model assessment – Insulin Resistance. HOMA-β: Homeostatic model assessment – β cell function. CIMT: carotid intima-media thickness. FM: whole-body fat mass. HDL-chol, legs FM | total FM and appendicular FM | truncal FM are the multiplicative inverse (1/x).

**Table S6** Epigenetic-age-related phenotyping of ObAGE participants by BMI trajectory (n=205)

| | TG1 (n=89) | TG2 (n=43) | TG (n=73) | Between-group Differences* | Cohen's f |
|---|---|---|---|---|---|
| Chronological age (y) | 29.1 ± 0.4 | 28.7 ± 0.4 | 28.8 ± 0.4 | NS | … |
| DNAm TL (kb) | 8.01 ± 0.3 | 7.46 ± 0.3 | 7.42 ± 0.2 | ABB | 0.77 (0.63-0.92) (S) |
| DNAm age Horvath (y) | 28.5 ± 2.5 | 33.1 ± 3.7 | 33.5 ± 4.3 | ABB | 0.69 (0.54-0.80) (S) |
| DNAm age Horvath acceleration (y) | -0.6 ± 2.5 | 4.3 ± 3.7 | 4.7 ± 4.2 | ABB | 0.73 (0.60-0.86) (S) |
| DNAm age Horvath acceleration (%) | -1.9 ± 8.6 | 15.2 ± 13.2 | 16.4 ± 14.1 | ABB | 0.73 (0.60-0.86) (S) |
| DNAm GrimAge (y) | 26.1 ± 3.5 | 31.0 ± 3.7 | 31.9 ± 3.9 | ABB | 0.68 (0.55-0.83) (S) |
| DNAm GrimAge acceleration (y) | -3.0 ± 3.5 | 2.2 ± 3.6 | 3.1 ± 3.8 | ABB | 0.76 (0.62-0.90) (S) |
| DNAm GrimAge acceleration (%) | -10.6 ± 1.2 | 7.7 ± 1.3 | 10.7 ± 1.3 | ABB | 0.76 (0.62-0.90) (S) |

Values expressed as mean ± SD, *HSD Tukey post-hoc test: A=TG1; B=TG2; C=TG3. Different letters indicate significant statistical differences. The same letter denotes means that do not differ. The effect size for the difference was computed as Cohen's *f* coefficient: 0.10 = weak effect size, 0.25 = moderate effect size, and 0.40 = strong effect size. TG1 refers to participants always having a BMI in the healthy range; TG2 refers to participants with obesity starting in adolescence and remaining obese into adulthood; and TG3 refers to participants who were obese in early childhood and remained obese into adulthood (TG3). TL: Telomere length.

**Table S7 Low-grade inflammation phenotyping of ObAGE participants by BMI trajectory (n=205)**

| | TG1 (n=89) | TG2 (n=43) | TG (n=73) | Between-group Differences* | Cohen's f |
|---|---|---|---|---|---|
| hs C Reactive Protein (mg/l) | 1.69 ± 2.1 | 3.67 ± 2.8 | 4.24 ± 2.4 | ABB | 0.41 (S) |
| Interleukin 2 (log) | 0.41 ± 0.3 | 0.66 ± 0.4 | 0.65 ± 0.5 | ABB | 0.29 (M) |
| Interleukin 6 (log) | 0.69± 0.5 | 1.03 ± 0.4 | 0.99 ± 0.4 | ABB | 0.46 (S) |
| Interleukin 10 (log) | 2.18 ± 0.3 | 2.27 ± 0.3 | 2.36 ± 0.3 | ABB | 0.27 (M) |
| Tumoral Necrosis Factor α (log) | 2.49 ± 0.5 | 2.52 ± 0.5 | 2.63 ± 0.6 | … | … |
| Fibroblast growth factor 21 (log) | 2.21 ± 0.2 | 2.42 ± 0.2 | 2.45 ± 0.2 | ABB | 0.54 (S) |
| Growth differentiation factor 11 (log) | 1.22 ± 0.05 | 1.23 ± 0.05 | 1.23 ± 0.05 | … | … |
| Growth differentiation factor 15 (log) | 3.44 ± 0.1 | 3.45 ± 0.1 | 3.51 ± 0.1 | AAC | 0.31 (M) |
| Insulin-like growth factor 1 | 4.65 ± 0.1 | 4.55 ± 0.1 | 4.45 ± 0.2 | ABC | 0.55 (S) |
| Insulin-like growth factor 2 | 5.54 ± 0.08 | 5.46 ± 0.09 | 5.44 ± 0.1 | ABB | 0.46 (S) |
| Leptin males (log) | 3.81 ± 0.4 | 4.28 ± 0.2 | 4.31 ± 0.2 | ABB | 0.77 (S) |
| Leptin females (log) | 4.23 ± 0.3 | 4.66 ± 0.3 | 4.75 ± 0.3 | ABB | 0.75 (S) |
| CD4 \| CD8 ratio | 1.78 (0.78) | 1.47 (0.52) | 1.48 (0.74) | ABB** | 0.02 (W) |
| Apelin (log) | 5.49 ± 0.1 | 5.65 ± 0.2 | 5.63 ± 0.2 | ABB | 0.41 (S) |
| Myostatin (log) | 7.06 ± 0.2 | 7.19 ± 0.2 | 7.16 ± 0.2 | ABB | 0.25 (M) |
| Irisin (log) | 7.71 ± 0.1 | 7.83 ± 0.2 | 7.80 ± 0.2 | ABB | 0.29 (M) |
| Oncostatin (log) | 1.59 ± 0.2 | 1.72 ± 0.2 | 1.70 ±0.2 | ABB | 0.32 (M) |
| Musclin (log) | 6.21 ± 0.2 | 6.08 ± 0.1 | 6.04 ± 0.1 | ABB | 0.51 (S) |
| Osteonectin (log) | 5.18 ± 0.2 | 5.27 ± 0.2 | 5.34 ± 0.2 | ABB | 0.32 (M) |

Values are expressed as mean ± SD or median (IQR). * HSD Tukey post-hoc test: A=TG1; B=TG2; C=TG3. Different letters indicate significant statistical differences. The same letter denotes means that do not differ. The ES for the ANOVA test was computed as Cohen's *f* coefficient: 0.10 = weak effect size, 0.25 = moderate effect size, and 0.40 = strong effect size. TG1: participants always having a BMI in the healthy range; TG2: participants with obesity starting in adolescence and remaining obese into adulthood; TG3: participants who were obese in early childhood and remaining obese into adulthood. ** The effect size for the Kruskal-Wallis test is computed as the ε2 based on the H-statistic: ε2 [H] = (H - k + 1)/(n - k), where H is the value obtained in the Kruskal-Wallis test; k is the number of groups; n is the total number of observations. The Dunn post hoc test was conducted to determine between-group differences: A=TG1; B=TG2; C=TG3, with different letters indicating significant statistical differences and the same letter denoting medians that do not differ. The ε2 estimate assumes values from 0 to 1; multiplied by 100, it indicates the percentage of variance in the dependent variable explained by the independent variable. The interpretation values commonly in published literature are 0.01- < 0.06 (weak effect), 0.06 - < 0.14 (moderate effect), and >= 0.14 (strong effect).

**Table S8 Adjacency matrix in males: molecular aging signatures at 29y old assessment**

| | DNAm age | DNAm TL | hs CRP | IL2 | IL6 | CD4\|CD8 | IL10 | Insulin | IGF1 | IGF2 | Apelin | Myostatin | Oncostatin | Irisin | Osteonectin | Osteocrin | FGF21 | GDF15 | Leptin | TNFa | GDF11 |
|---|---|---|---|---|---|---|---|---|---|---|---|---|---|---|---|---|---|---|---|---|---|
| DNAm age | **0** | 0.027 | 0.239 | 0.144 | 0.337 | 0.037 | 1.196 | 0.231 | 0.154 | 0.059 | 1.190 | 0.985 | 0.781 | 1.187 | 0.518 | 1.492 | 0.089 | 0.187 | 0.256 | 0.155 | 0.031 |
| DNAm TL | 0.027 | **0** | 0.219 | 0.122 | 0.318 | 0.012 | 1.184 | 0.210 | 0.132 | 0.035 | 1.177 | 0.972 | 0.766 | 1.175 | 0.501 | 1.481 | 0.065 | 0.166 | 0.236 | 0.133 | 0.003 |
| hs CRP | 0.239 | 0.219 | **0** | 0.014 | 0.010 | 0.131 | 0.411 | 0.000 | 0.011 | 0.080 | 0.407 | 0.285 | 0.175 | 0.406 | 0.060 | 0.605 | 0.046 | 0.004 | 0.000 | 0.011 | 0.271 |
| IL2 | 0.144 | 0.122 | 0.014 | **0** | 0.047 | 0.059 | 0.574 | 0.012 | 0.000 | 0.027 | 0.569 | 0.424 | 0.288 | 0.567 | 0.133 | 0.797 | 0.009 | 0.003 | 0.019 | 0.000 | 0.161 |
| IL6 | 0.337 | 0.318 | 0.010 | 0.047 | **0** | 0.210 | 0.298 | 0.011 | 0.042 | 0.145 | 0.294 | 0.191 | 0.103 | 0.293 | 0.022 | 0.467 | 0.097 | 0.025 | 0.006 | 0.041 | 0.379 |
| CD4CD8 | 0.037 | 0.012 | 0.131 | 0.059 | 0.210 | **0** | 0.976 | 0.124 | 0.066 | 0.006 | 0.970 | 0.782 | 0.597 | 0.968 | 0.364 | 1.253 | 0.022 | 0.090 | 0.144 | 0.067 | 0.026 |
| IL10 | 1.196 | 1.184 | 0.411 | 0.574 | 0.298 | 0.976 | **0** | 0.423 | 0.554 | 0.835 | 0.000 | 0.012 | 0.051 | 0.000 | 0.160 | 0.020 | 0.719 | 0.491 | 0.388 | 0.550 | 1.293 |
| Insulin | 0.231 | 0.210 | 0.000 | 0.012 | 0.011 | 0.124 | 0.423 | **0** | 0.009 | 0.075 | 0.419 | 0.295 | 0.183 | 0.417 | 0.065 | 0.619 | 0.042 | 0.003 | 0.001 | 0.009 | 0.261 |
| IGF1 | 0.154 | 0.132 | 0.011 | 0.000 | 0.042 | 0.066 | 0.554 | 0.009 | **0** | 0.031 | 0.549 | 0.407 | 0.274 | 0.547 | 0.123 | 0.774 | 0.012 | 0.002 | 0.016 | 0.000 | 0.173 |
| IGF2 | 0.059 | 0.035 | 0.080 | 0.027 | 0.145 | 0.006 | 0.835 | 0.075 | 0.031 | **0** | 0.829 | 0.655 | 0.485 | 0.827 | 0.277 | 1.095 | 0.005 | 0.049 | 0.091 | 0.032 | 0.057 |
| Apelin | 1.190 | 1.177 | 0.407 | 0.569 | 0.294 | 0.970 | 0.000 | 0.419 | 0.549 | 0.829 | **0** | 0.011 | 0.050 | 0.000 | 0.157 | 0.021 | 0.713 | 0.487 | 0.384 | 0.545 | 1.286 |
| Myostatin | 0.985 | 0.972 | 0.285 | 0.424 | 0.191 | 0.782 | 0.012 | 0.295 | 0.407 | 0.655 | 0.011 | **0** | 0.014 | 0.011 | 0.085 | 0.063 | 0.551 | 0.353 | 0.266 | 0.403 | 1.072 |
| Oncostatin | 0.781 | 0.766 | 0.175 | 0.288 | 0.103 | 0.597 | 0.051 | 0.183 | 0.274 | 0.485 | 0.050 | 0.014 | **0** | 0.049 | 0.030 | 0.135 | 0.395 | 0.229 | 0.160 | 0.271 | 0.857 |
| Irisin | 1.187 | 1.175 | 0.406 | 0.567 | 0.293 | 0.968 | 0.000 | 0.417 | 0.547 | 0.827 | 0.000 | 0.011 | 0.049 | **0** | 0.156 | 0.021 | 0.711 | 0.485 | 0.383 | 0.544 | 1.283 |
| Osteonectin | 0.518 | 0.501 | 0.060 | 0.133 | 0.022 | 0.364 | 0.160 | 0.065 | 0.123 | 0.277 | 0.157 | 0.085 | 0.030 | 0.156 | **0** | 0.291 | 0.210 | 0.094 | 0.051 | 0.121 | 0.577 |
| Osteocrin | 1.492 | 1.481 | 0.605 | 0.797 | 0.467 | 1.253 | 0.020 | 0.619 | 0.774 | 1.095 | 0.021 | 0.063 | 0.135 | 0.021 | 0.291 | **0** | 0.963 | 0.700 | 0.578 | 0.769 | 1.600 |
| FGF21 | 0.089 | 0.065 | 0.046 | 0.009 | 0.097 | 0.022 | 0.719 | 0.042 | 0.012 | 0.005 | 0.713 | 0.551 | 0.395 | 0.711 | 0.210 | 0.963 | **0** | 0.023 | 0.054 | 0.012 | 0.095 |
| GDF15 | 0.187 | 0.166 | 0.004 | 0.003 | 0.025 | 0.090 | 0.491 | 0.003 | 0.002 | 0.049 | 0.487 | 0.353 | 0.229 | 0.485 | 0.094 | 0.700 | 0.023 | **0** | 0.006 | 0.002 | 0.211 |
| Leptin | 0.256 | 0.236 | 0.000 | 0.019 | 0.006 | 0.144 | 0.388 | 0.001 | 0.016 | 0.091 | 0.384 | 0.266 | 0.160 | 0.383 | 0.051 | 0.578 | 0.054 | 0.006 | **0** | 0.015 | 0.290 |
| TNFa | 0.155 | 0.133 | 0.011 | 0.000 | 0.041 | 0.067 | 0.550 | 0.009 | 0.000 | 0.032 | 0.545 | 0.403 | 0.271 | 0.544 | 0.121 | 0.769 | 0.012 | 0.002 | 0.015 | **0** | 0.175 |
| GDF11 | 0.031 | 0.003 | 0.271 | 0.161 | 0.379 | 0.026 | 1.293 | 0.261 | 0.173 | 0.057 | 1.286 | 1.072 | 0.857 | 1.283 | 0.577 | 1.600 | 0.095 | 0.211 | 0.290 | 0.175 | **0** |

Hierarchical cluster analysis on standardized data of selected age-related biomarkers assessed at 28- 29y. Distance criteria: Euclidian method. Linkage criteria: Ward linkage. hs CRP: high sensitivity C reactive protein. TL: telomere length. IL2: interleukin 2. IL6: interleukin 6. IL10: interleukin 10. IGF-1: insulin-like growth factor 1. IGF-2: insulin-like growth factor 2. TNF α: Tumoral Necrosis Factor α. GDF11: Growth differentiation factor 11. GDF15: Growth differentiation factor 15. FGF21: Fibroblast growth factor 21. Variables were standardized for analysis purposes. DNAmTL, IGF1, IGF2, Osteocrin (Musclin), and CD4|CD8 ratio were expressed as the multiplicative inverse (1/x) for analysis.

**Table S9 Adjacency matrix in females: molecular aging signatures at 29y old assessment**

| | DNAm age | DNAm TL | hs CRP | IL2 | IL6 | CD4CD8 | IL10 | Insulin | IGF1 | IGF2 | Apelin | Myostatin | Oncostatin | Irisin | Osteonectin | Osteocrin | FGF21 | GDF15 | Leptin | TNFa | GDF11 |
|---|---|---|---|---|---|---|---|---|---|---|---|---|---|---|---|---|---|---|---|---|---|
| DNAm age | **0** | 0.005 | 0.000 | 0.770 | 0.000 | 1.540 | 0.978 | 0.382 | 0.057 | 0.163 | 0.039 | 0.580 | 0.008 | 0.003 | 0.003 | 1.524 | 0.632 | 0.005 | 0.206 | 0.632 | 0.044 |
| DNAm TL* | 0.005 | **0** | 0.005 | 0.652 | 0.007 | 1.706 | 0.847 | 0.299 | 0.027 | 0.110 | 0.016 | 0.477 | 0.026 | 0.015 | 0.015 | 1.364 | 0.524 | 0.000 | 0.146 | 0.525 | 0.018 |
| hs CRP | 0.000 | 0.005 | **0** | 0.765 | 0.000 | 1.546 | 0.973 | 0.379 | 0.056 | 0.161 | 0.038 | 0.576 | 0.008 | 0.003 | 0.003 | 1.518 | 0.627 | 0.004 | 0.204 | 0.628 | 0.042 |
| IL2 | 0.770 | 0.652 | 0.765 | **0** | 0.787 | 3.874 | 0.014 | 0.072 | 0.419 | 0.235 | 0.472 | 0.015 | 0.925 | 0.857 | 0.857 | 0.148 | 0.007 | 0.660 | 0.189 | 0.007 | 0.458 |
| IL6 | 0.000 | 0.007 | 0.000 | 0.787 | **0** | 1.517 | 0.998 | 0.395 | 0.062 | 0.172 | 0.043 | 0.595 | 0.006 | 0.002 | 0.002 | 1.547 | 0.648 | 0.006 | 0.216 | 0.648 | 0.048 |
| CD4\|CD8 | 1.540 | 1.706 | 1.546 | 3.874 | 1.517 | **0** | 4.208 | 3.120 | 2.105 | 2.532 | 2.004 | 3.532 | 1.346 | 1.428 | 1.428 | 4.954 | 3.629 | 1.695 | 2.667 | 3.630 | 2.031 |
| IL10 | 0.978 | 0.847 | 0.973 | 0.014 | 0.998 | 4.208 | **0** | 0.149 | 0.581 | 0.361 | 0.642 | 0.057 | 1.150 | 1.075 | 1.075 | 0.071 | 0.042 | 0.855 | 0.303 | 0.042 | 0.625 |
| Insulin | 0.382 | 0.299 | 0.379 | 0.072 | 0.395 | 3.120 | 0.149 | **0** | 0.147 | 0.047 | 0.179 | 0.022 | 0.497 | 0.446 | 0.446 | 0.420 | 0.033 | 0.304 | 0.028 | 0.033 | 0.170 |
| IGF1 | 0.057 | 0.027 | 0.056 | 0.419 | 0.062 | 2.105 | 0.581 | 0.147 | **0** | 0.028 | 0.002 | 0.280 | 0.107 | 0.084 | 0.084 | 1.031 | 0.317 | 0.029 | 0.047 | 0.317 | 0.001 |
| IGF2 | 0.163 | 0.110 | 0.161 | 0.235 | 0.172 | 2.532 | 0.361 | 0.047 | 0.028 | **0** | 0.043 | 0.133 | 0.242 | 0.206 | 0.206 | 0.737 | 0.159 | 0.113 | 0.003 | 0.159 | 0.038 |
| Apelin | 0.039 | 0.016 | 0.038 | 0.472 | 0.043 | 2.004 | 0.642 | 0.179 | 0.002 | 0.043 | **0** | 0.324 | 0.082 | 0.062 | 0.062 | 1.110 | 0.364 | 0.017 | 0.067 | 0.364 | 0.000 |
| Myostatin | 0.580 | 0.477 | 0.576 | 0.015 | 0.595 | 3.532 | 0.057 | 0.022 | 0.280 | 0.133 | 0.324 | **0** | 0.718 | 0.657 | 0.657 | 0.254 | 0.001 | 0.484 | 0.099 | 0.001 | 0.312 |
| Oncostatin | 0.008 | 0.026 | 0.008 | 0.925 | 0.006 | 1.346 | 1.150 | 0.497 | 0.107 | 0.242 | 0.082 | 0.718 | **0** | 0.001 | 0.001 | 1.727 | 0.775 | 0.025 | 0.294 | 0.775 | 0.089 |
| Irisin | 0.003 | 0.015 | 0.003 | 0.857 | 0.002 | 1.428 | 1.075 | 0.446 | 0.084 | 0.206 | 0.062 | 0.657 | 0.001 | **0** | 0.000 | 1.639 | 0.712 | 0.014 | 0.255 | 0.712 | 0.067 |
| Osteonectin | 0.003 | 0.015 | 0.003 | 0.857 | 0.002 | 1.428 | 1.075 | 0.446 | 0.084 | 0.206 | 0.062 | 0.657 | 0.001 | 0.000 | **0** | 1.639 | 0.712 | 0.014 | 0.255 | 0.712 | 0.067 |
| Osteocrin | 1.524 | 1.364 | 1.518 | 0.148 | 1.547 | 4.954 | 0.071 | 0.420 | 1.031 | 0.737 | 1.110 | 0.254 | 1.727 | 1.639 | 1.639 | **0** | 0.221 | 1.375 | 0.656 | 0.220 | 1.088 |
| FGF21 | 0.632 | 0.524 | 0.627 | 0.007 | 0.648 | 3.629 | 0.042 | 0.033 | 0.317 | 0.159 | 0.364 | 0.001 | 0.775 | 0.712 | 0.712 | 0.221 | **0** | 0.532 | 0.121 | 0.000 | 0.351 |
| GDF15 | 0.005 | 0.000 | 0.004 | 0.660 | 0.006 | 1.695 | 0.855 | 0.304 | 0.029 | 0.113 | 0.017 | 0.484 | 0.025 | 0.014 | 0.014 | 1.375 | 0.532 | **0** | 0.150 | 0.532 | 0.020 |
| Leptin | 0.206 | 0.146 | 0.204 | 0.189 | 0.216 | 2.667 | 0.303 | 0.028 | 0.047 | 0.003 | 0.067 | 0.099 | 0.294 | 0.255 | 0.255 | 0.656 | 0.121 | 0.150 | **0** | 0.122 | 0.061 |
| TNFa | 0.632 | 0.525 | 0.628 | 0.007 | 0.648 | 3.630 | 0.042 | 0.033 | 0.317 | 0.159 | 0.364 | 0.001 | 0.775 | 0.712 | 0.712 | 0.220 | 0.000 | 0.532 | 0.122 | **0** | 0.351 |
| GDF11 | 0.044 | 0.018 | 0.042 | 0.458 | 0.048 | 2.031 | 0.625 | 0.170 | 0.001 | 0.038 | 0.000 | 0.312 | 0.089 | 0.067 | 0.067 | 1.088 | 0.351 | 0.020 | 0.061 | 0.351 | **0** |

Hierarchical cluster analysis on standardized data of selected age-related biomarkers assessed at 28- 29y. Distance criteria: Euclidian method. Linkage criteria: Ward linkage. hs CRP: high sensitivity C reactive protein. TL: telomere length. IL2: interleukin 2. IL6: interleukin 6. IL10: interleukin 10. IGF-1: insulin-like growth factor 1. IGF-2: insulin-like growth factor 2. TNF α: Tumoral Necrosis Factor α. GDF11: Growth differentiation factor 11. GDF15: Growth differentiation factor 15. FGF21: Fibroblast growth factor 21. Variables were standardized for analysis purposes. DNAmTL, IGF1, IGF2, Osteocrin (Musclin), and CD4|CD8 ratio were expressed as the multiplicative inverse (1/x) for analysis.

**Table S10 Summary of body composition markers in the resilient subject compared to the body composition profile in TG3 and TG1**

| | Subject A | Subject B | Subject C | Subject D | Subject E | Subject F | Subject G | TG3 | TG1 |
|---|---|---|---|---|---|---|---|---|---|
| Body mass Index | 30.10 | 43.40 | 47.90 | 43.00 | 42.10 | 34.90 | 41.00 | 37.70 | 23.10 |
| Lean mass (%) | 54.10 | 48.70 | 42.70 | 41.40 | 44.70 | 58.00 | 49.30 | 51.00 | 63.00 |
| Fat mass (%) | 42.80 | 49.00 | 55.50 | 56.20 | 52.70 | 38.70 | 48.00 | 45.00 | 33.00 |
| Truncal fat mass (%) | 32.30 | 39.20 | 45.80 | 36.10 | 37.50 | 36.80 | 40.40 | 37.60 | 27.00 |
| Waist circumference (cm) | 85.10 | 114.50 | 126.70 | 105.00 | 103.40 | 119.10 | 104.00 | 109.00 | 76.00 |
| Waist-to-Height ratio | 53.52 | 72.93 | 80.19 | 66.04 | 65.86 | 66.91 | 65.00 | 66.00 | 46.00 |

**Table S11 Summary of aging biomarkers in the resilient subject compared to the profile in TG3 and TG1**

| | Subject A | Subject B | Subject C | Subject D | Subject E | Subject F | Subject G | TG1 | TG3 |
|---|---|---|---|---|---|---|---|---|---|
| DNAm Age (y) | 26.70 | 27.10 | 23.50 | 23.60 | 26.00 | 25.50 | 26.20 | 28.50 | 34.00 |
| DNAm GrimAge (y) | 28.50 | 26.50 | 26.50 | 24.20 | 28.30 | 26.20 | 26.30 | 26.00 | 32.00 |
| hs CRP (mg/l) | 2.40 | 1.50 | **5.70** | **8.90** | **7.00** | **4.90** | **3.00** | 1.69 | **4.24** |
| CD4 \| CD8 ratio | 1.20 | 1.40 | 1.65 | 1.64 | 1.72 | 1.16 | 1.78 | 1.78 | 1.47 |
| DNAm TL (kb) | 7.57 | 7.37 | 7.25 | 7.51 | 7.53 | 7.54 | 7.52 | 8.10 | 7.42 |
| Apelin (log) | 5.56 | 5.63 | 5.74 | 5.75 | 5.96 | 5.45 | 5.65 | 5.49 | 5.63 |
| Musclin (log) | 6.13 | 6.08 | 5.94 | 6.15 | 6.17 | 5.92 | 6.10 | 6.21 | 6.04 |
| IL-6 (log) | 1.49 | 0.94 | 0.43 | 0.93 | 0.30 | 0.67 | 1.66 | 0.69 | 0.99 |
| IGF-1 (log) | 4.65 | 4.61 | 4.63 | 4.66 | 4.51 | 4.52 | 4.62 | 4.65 | 4.45 |
| IGF-2 (log) | 5.52 | 5.54 | 5.51 | 5.58 | 5.53 | 5.62 | 5.52 | 5.54 | 5.44 |

hs CRP: high sensitivity C reactive protein. TL: telomere length. IL6: interleukin 6. IGF-1: insulin-like growth factor 1. IGF-2: insulin-like growth factor 2

Table S12 Summary of cardiometabolic markers in the resilient subjects compared to the cardiometabolic profile in TG3 and TG1

| | **Subject A** | **Subject B** | **Subject C** | **Subject D** | **Subject E** | **Subject F** | **Subject G** | **TG3** | **TG1** |
|---|---|---|---|---|---|---|---|---|---|
| Waist circumference (cm) | 85.10 | 114.50 | 126.70 | 105.00 | 103.40 | 119.10 | 104.00 | 107.1 | 75.6 |
| Systolic BP (mmHg) | 117 | 131 | 130 | 125 | 118 | 142 | 118 | 127 | 118 |
| Diastolic BP (mmHg) | 75 | 74 | 80 | 78 | 83 | 83 | 77 | 77 | 75 |
| Glycemia (mg/dl) | 83.1 | 99.9 | 104.2 | 93.6 | 83.3 | 102.3 | 96.9 | 97.6 | 88.0 |
| Triglycerides (mg/dl) | 38.0 | 71.0 | 221.9 | 92.6 | 139.1 | 285.0 | 129.1 | 121.0 | 84.00 |
| HDL cholesterol (mg/dl) | 30.3 | 17.3 | 23.5 | 41.6 | 18.6 | 10.9 | 13.8 | 19.0 | 36.0 |
| LDL cholesterol (mg/dl) | 77.5 | 163.4 | 140.0 | 74.2 | 130.9 | 72.6 | 125.9 | 147.0 | 109.0 |
| Insulin (uU/l) | 9.3 | 18.9 | 23.7 | 13.2 | 22.9 | 17.7 | 39.1 | 17.0 | 10.4 |
| HOMA-IR | 1.90 | 4.70 | 6.10 | 3.00 | 4.70 | 4.50 | 9.30 | 4.10 | 2.10 |
| CDM risk factors (n) | 2 | 4 | 4 | 2 | 2 | 5 | 2 | **…** | … |

BP: blood pressure. HDL: high-density lipoprotein. LDL: low-density lipoprotein. HOMA-IR: Homeostatic model assessment – Insulin Resistance. CDM risk factors: cardiometabolic risk factors (0-5).
